# Estimating the impact of *Shigella* vaccines on growth outcomes and implications for clinical trial design

**DOI:** 10.64898/2026.04.03.26350105

**Authors:** Allison Codi, Elizabeth Rogawski McQuade, David Benkeser

**Author notes:** Corresponding author Email address (Allison Codi).

## Abstract

**Background:** The value proposition for *Shigella* vaccines is strengthened by the potential for vaccines to prevent linear growth faltering. However, because expected effect sizes in Phase 3 vaccine trials are small due to limited Shigella incidence, a simple comparison of growth by randomized vaccine arm is likely underpowered and may yield null or even inverse results.

**Methods:** We consider a new approach that estimates vaccine effects in the subgroup that would be infected in absence of vaccination, termed the *naturally infected*. In simulations parameterized by multi-site studies of diarrhea, we compare power for detecting linear growth effects in the naturally infected versus the full study. We further quantified how power is impacted by trial design choices including immunization schedule, study site, and timing of growth measurements.

**Findings:** Simple comparisons of height-for-age z-score (HAZ) by randomized vaccine arm have extremely limited power (<15%) at realistic trial sizes (n=2,500 to 20,000) and carry risk of showing an inverse effect due to random chance. In contrast, naturally infected effects were five to ten times larger and power was up to three times higher. Using a twelve month immunization schedule with a single growth endpoint in high-incidence settings maximized power to detect an effect.

**Interpretations:** While realistically sized clinical trials may be underpowered to detect an effect of vaccination on growth, estimation using the naturally infected subpopulation and careful trial design improve chances of detecting an effect while mitigating risks of null or inverse results.

**Funding:** This work was supported by the Gates Foundation (INV-071320 to ERM and DB).

## Introduction

*Shigella* is a leading cause of acute diarrheal disease among young children in low- and middle-income settings,^1, 2^ with 30% experiencing *Shigella*-attributable diarrhea by age two.^3^ It ranks as the second leading cause of diarrheal deaths among children under five with an estimated 64,000 deaths annually.^4^ Beyond acute morbidity and mortality, several large, multi-site studies have shown that *Shigella* infection is associated with long-term linear growth faltering.^1, 5^ Poor growth in the first two years of life increases risk of chronic disease, lower educational attainment, and limited economic success lasting into adulthood.^6^ While *Shigella* can be treated effectively with antibiotics, growing concern of antimicrobial resistance^7^ has made the development of *Shigella* vaccines a growing priority.

Several candidate *Shigella* vaccines are in development,^8^ including bivalent and quadrivalent formulations that are entering Phase 2/3 clinical trials (e.g., NCT06838195). While the primary endpoint of these trials is likely to be *Shigella*-attributed moderate-to-severe diarrhea,^9^ measuring the effect of the vaccine on linear growth faltering is key to understanding the full value of a vaccine. Modeling studies suggest that even small effects of the vaccine on linear growth can have beneficial public health and economic impacts.^10, 11^ Accordingly, the World Health Organization (WHO) has identified reduction in height-for-age z-score (HAZ), an indicator of linear growth faltering, as a desired secondary endpoint in clinical trials.^9^

Detecting vaccine effects on post-infection outcomes such as growth faltering is challenging. Vaccine effects are traditionally estimated by comparing outcomes between those randomized to vaccine versus placebo. However, children who are not infected with *Shigella* are not at risk for *Shigella*-attributable growth faltering, so are unlikely to experience a vaccine effect on growth. As a result, a comparison of HAZ between trial arms will be largely diluted by those who remain uninfected. This dilution may substantially reduce the improvement in HAZ attributable to vaccination, reduce statistical power, and increase the likelihood of observing null or even spuriously harmful estimates. Alternatively, restricting the comparison of HAZ to children who become infected excludes children for whom the vaccine prevented *Shigella* (i.e., the children we expect to benefit most), and may introduce selection bias.^12^

In this study, we use a causal inference approach to estimate the effect of *Shigella* vaccination on HAZ among children who would have had *Shigella* diarrhea in the absence of vaccination. We have previously developed methods to evaluate vaccine effects in this subgroup, termed the *naturally infected*.^13^ We use a realistic simulation study to compare statistical power to detect vaccine effects in the naturally infected with power to detect effects in the full study population. Our simulation also explores how trial design features influence expected growth effect size and statistical power for detecting vaccine growth effects. Previous work suggests that the effect of *Shigella* infection on linear growth varies by age and baseline HAZ,^14^ such that study site selection and dose schedule may substantially impact expected growth effect sizes. Furthermore, the timing and frequency of growth measurements, considerations for trial design that have substantial resource implications, may affect both effect size and power. We explore the impact of these features in our simulation study.

This analysis provides guidance for trial design and statistical analysis for the evaluation of linear growth faltering as a secondary endpoint in *Shigella* vaccine trials.

## 1. Methods

### 1.1 Study design and analysis approach

We simulate 1:1 randomized, placebo-controlled trials with one year of follow-up under a range of study designs aligned with WHO Preferred Product Characteristics (PPC)^9^ and practical considerations for Phase 3 trial planning. In each simulated trial, the primary endpoint is defined as PCR-attributable moderate-to-severe *Shigella* diarrhea.^15^ The growth endpoint of interest is defined by height-for-age z-score measured at prespecified time points during follow-up. Children are enrolled at ages corresponding to each immunization schedule and randomized to receive vaccine or placebo, with relevant demographic and clinical characteristics collected at enrollment. Participants are followed for 12 months after achieving full vaccine protection, with active surveillance to capture all *Shigella*-attributable diarrhea (i.e., 100% medical attendance). Analyses are conducted under an intention-to-treat framework, and assume 10% study drop out.

We calculate sample size needed to achieve 90% power for the primary endpoint using Farrington and Manning score test for relative risk,^16^ with vaccine efficacy of 60% against moderate-to-severe disease and a lower 95% confidence interval bound of 20%, in line with previous calculations.^17^

We compare a population-level estimate of the effect of *Shigella* vaccination on HAZ to an approach that estimates the effect among children who would have had *Shigella*-attributable diarrhea in the absence of vaccination, referred to as the *naturally infected*. Figure 1 illustrates this concept using an example with 10 children. Under placebo, four children would experience *Shigella*-attributable diarrhea during follow-up, and six children would not. The vaccine would prevent two of these four children from having *Shigella* diarrhea. When HAZ is compared across all ten participants, i.e. a population-level approach, children who never had *Shigella* diarrhea are included but have the same HAZ under vaccine and placebo, thus diluting the effect. Restricting to those who would have had *Shigella* diarrhea under placebo leads to a larger effect size and better reflects the biological impact of vaccination on growth, which is likely of most interest in a Phase 3 trial.

**Figure 1:**
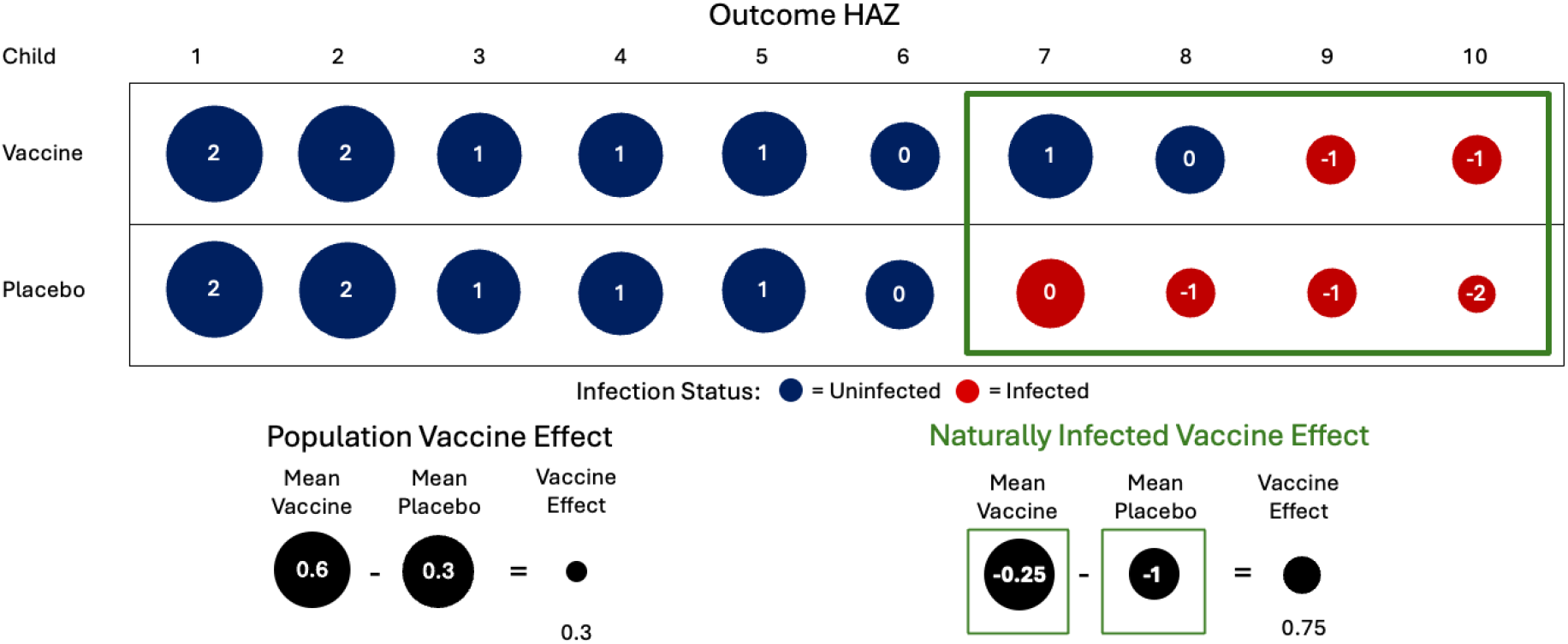
Illustration of population-level and naturally infected approaches. Each circle represents a child’s HAZ under vaccine or placebo. Blue circles denote children who remain *Shigella* diarrhea free, and red circles denote children who experience *Shigella*-attributable diarrhea. The green box identifies children who would have had *Shigella* diarrhea under placebo, corresponding to the naturally infected subset.

In practice, we cannot observe the *Shigella* status of a single child under both placebo and vaccine. Thus, the naturally infected approach requires several causal assumptions, which we argue are plausible in a vaccine trial setting. A key assumption is that baseline covariates influencing both growth faltering and the risk of *Shigella*-attributable diarrhea are controlled for. Our simulations are parameterized such that confounding operates through baseline HAZ and therefore we only adjust for this baseline covariate. In practice, a broader set of baseline demographic and clinical characteristics should be collected to satisfy this assumption. For details on the naturally infected approach and associated assumptions, see Supplementary Section 1.

### 1.2 Simulation Parameterization

Simulations were parameterized using data from multi-site studies of diarrhea that have shown an association between *Shigella* and linear growth faltering.^14^ We describe parameterization for a trial with an immunization schedule that confers complete protection by 6-months in a site with average incidence and average expected effect of *Shigella*-diarrhea on HAZ, which we term the *general recruitment scenario* (Figure 2). Parameters used in additional scenarios are listed in Supplement Section 2.

**Figure 2:**
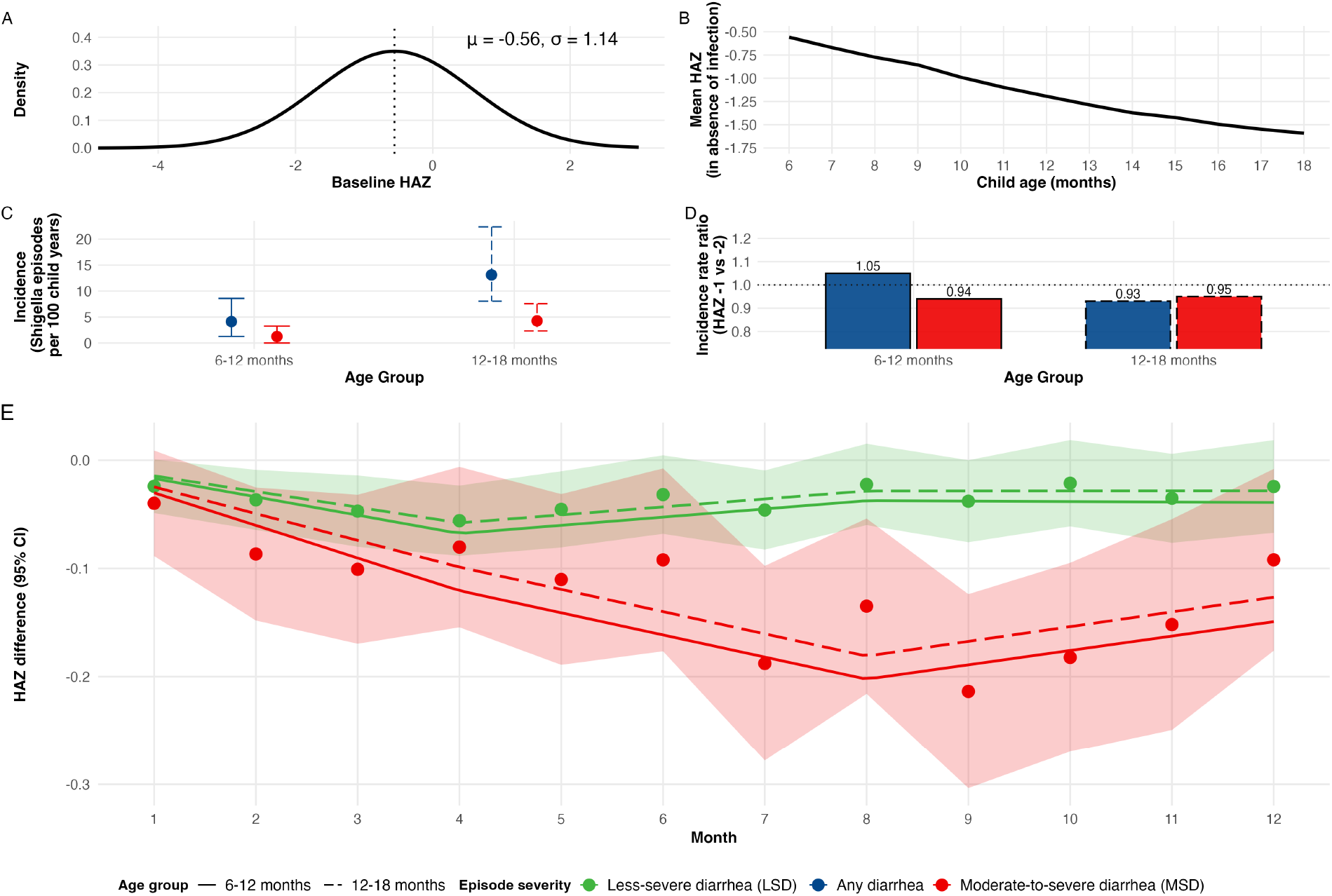
Simulation parameterization for vaccine trial in a general recruitment setting with immunization schedule completed by 6 months of age. (A) Distribution of baseline HAZ. (B) One-year average growth trajectory in absence of infection for children with mean baseline HAZ. (C) Average incidence of MAD and MSD in young children (first 6 months of follow up for a trial enrolling 6 month olds) and older children (second half of the trial). (D) Incidence was simulated to vary by HAZ. The incidence rate ratio for a one-unit increase of baseline HAZ from -2 to -1 for any *Shigella* diarrhea by age group is shown. (E) Monthly estimated effects of *Shigella* diarrhea by severity level (bold, dash). Splines were fit for each age strata separately, with younger children experiencing larger deleterious growth effects following *Shigella* diarrhea.

Baseline HAZ was simulated for each child from a Normal distribution with mean -0.56 and standard deviation 1.14 (Figure 2A). This closely mirrored the observed distribution of HAZ in children ages 6-8 months in The Gambia site of the Enterics for Global Health (EFGH) *Shigella* surveillance study, a hybrid surveillance study of medically attended diarrhea.^18^

Given baseline HAZ, one-year growth trajectories were simulated using monthly growth models fit to data from the Malnutrition and Enteric Diseases (MAL-ED) study, a prospective observational birth cohort.^1^ These models regressed HAZ at month *t* on HAZ at month *t −* 1 among children who had not experienced PCR-attributable *Shigella* diarrhea up to time *t*. Estimated parameters from these models were used to simulate HAZ trajectories that would be observed in the absence of *Shigella* diarrhea (see example trajectory, Figure 2B).

The Gambia EFGH data were used to estimate the marginal incidence of diarrhea attributable to *Shigella* serotypes *S. flexneri* 2a, 3a, and 6 or *S. sonnei*, the serotypes targeted by some quadrivalent vaccine candidates.^8, 19, 20^ Incidence of any *Shigella*-diarrhea and of moderate-to-severe (MSD) *Shigella* diarrhea were 4.2 and 2.9 episodes per 100 child-years among children ages 6-12 months, and 13.1 and 4.1 episodes per 100 child-years among children ages 12-18 months, respectively (Figure 2C).

To generate simulated timing of Shigella infections given baseline HAZ, we first fit Cox proportional hazards models to the MAL-ED data. Hazards given baseline HAZ were modeled separately for any diarrhea in the age intervals 6-12 months and 12-18 months. We then fit models for probability of a diarrhea episode being severe given baseline HAZ within EFGH data. These models showed a modestly protective association between higher HAZ and *Shigella* MSD from 6-18 months.

Growth trajectories of children who experienced *Shigella*-attributable diarrhea during the simulated trial were decremented based on estimates derived from MAL-ED. We estimated the effects of less-severe *Shigella* diarrhea (LSD) and *Shigella* MSD on linear growth every month for one year following the episode, adjusting for baseline covariates, severity characteristics, other pathogens detected, and antibiotic use (Figure 2E).^14^ We simulated growth decrements from these curves by fitting linear splines to the monthly estimates with knots at 4 and 8 months. Because prior evidence supports that younger children have larger growth impacts from *Shigella* infection,^5, 14^ we simulated larger growth decrements following infection in children aged 6-12 months (solid lines, Figure 2E) compared to children aged 12-18 months (dashed lines).

We assumed vaccine efficacy of 40% against LSD and 60% against MSD for all simulations, aligning with target efficacy in the WHO PPC.^9^

### 1.2.1 Study design choices

Several of the parameters described above are influenced by design choices in the Phase 3 trial. We consider the following three design characteristics in alternative trial scenarios: (i) immunization schedule, (ii) choice of study site, and (iii) timing and frequency of outcome growth measurements.

#### (i) Immunization schedule

The WHO PPC for a *Shigella* vaccine recommend vaccination as early as 6 months of age and within the first year of life.^9^ We therefore consider immunization schedules that provide complete protection by either 6 or 12 months of age, with follow-up spanning ages 6-18 months or 12-24 months, respectively. Immunization schedule influences *Shigella* diarrhea incidence and distribution of baseline HAZ in the simulated trials.

#### (ii) Recruitment strategy

To reflect heterogeneity in potential trial sites by age-specific *Shigella* incidence,^18^ we consider three recruitment strategies representing distinct epidemiologic settings. The first strategy, termed *general recruitment*, reflects a trial in a setting with moderately high incidence without any targeting of recruitment. We use point estimates of incidence from The Gambia EFGH site and use the average effect of *Shigella*-diarrhea on HAZ from MAL-ED. To reflect effect modification by age, we use splines fit to the growth effect point estimates and shift estimates upward by 25% within their confidence intervals for older children (18-24 months) and downward by 25% for younger children (6-12 months).

The second strategy, termed *targeted recruitment*, reflects enrollment restricted to children at highest risk of *Shigella*-attributable diarrhea and subsequent growth faltering. Practically, this could correspond to incorporating a-priori risk factors into eligibility criteria. We parametrize this scenario by assuming a higher incidence, reflected by use of the upper 95% confidence bound of incidence estimates from The Gambia in EFGH, and larger magnitude of growth effects, implemented by scaling HAZ estimates toward the lower 95% confidence bound of growth effect estimates from MAL-ED in all age groups. This yields a setting with both higher incidence and larger expected growth deficits within the targeted subpopulation.

The third strategy, *high early incidence*, targets settings with high incidence in the youngest age group (ages 6-12 months) relative to older children (particularly, ages 12-18 months). Younger children experience larger effects of *Shigella*-attributable diarrhea on HAZ, so higher incidence at younger ages may improve power for earlier immunization schedules. We parameterize this scenario using data from Peru in EFGH, where the ratio of incidence among children aged 6-12 months (13 MSD episodes per 100 child years) relative to that among ages 12-18 months (14 MSD episodes per 100 child years) is higher than all other EFGH sites.^18^ Growth effects are parametrized as in the general recruitment strategy.

The combination of design characteristics (i) and (ii) yields six trial scenarios–two immunization schedules (complete protection by 6 versus 12 months) with three recruitment strategies (general recruitment, targeted recruitment, high early incidence).

#### (iii) Timing and frequency of growth measurements

The timing and frequency of growth measurements may impact vaccine effect size since effects on HAZ differ based on age and time since *Shigella* infection. Measuring growth too soon after infection may yield smaller effect sizes, while waiting too long to measure growth may also limit effect sizes if catch-up growth occurs after initial faltering. Collecting interim growth measurements throughout the trial may improve power to detect effects, with the trade offs of increased trial costs and logistical complexities. We therefore compare simulated trials that evaluate the growth outcome mid-study (i.e., at 6 months of follow-up) with trials that evaluate a final growth measurement at 12 months of follow-up, and trials that make use of both measurements, in which case we consider the average of the effects at the two time points. In a sensitivity analysis, we consider trials that incorporate four outcome growth measurements.

### 1.3 Statistical Analysis

We simulated 1,000 datasets of sizes *n* = [2500, 5000, 10000, 20000, 40000, 80000]. For each dataset, we estimate the effect of vaccination on growth outcomes in the naturally infected and in the full population using augmented inverse probability weighted (AIPW) estimators.^13^

We estimated power by calculating the proportion of the 1,000 simulated datasets where a two-sided test of the null hypothesis of no growth effect rejected the null at level 0.05. We also calculated the proportion of simulations in which the estimated effect was negative, representing the probability that the trial would incorrectly suggest the vaccine harms growth.

## 2. Results

### 2.1 Naturally Infected vs Population Vaccine Effect

Vaccine effects on linear growth in the naturally infected were 5-10 times larger than population-level effects across settings. In the general recruitment scenario (6 month immunization schedule), the population-level effect on linear growth at 12 months was 0.002 HAZ, compared to 0.027 HAZ among the naturally infected.

While power was suboptimal in almost all cases, power to detect the vaccine effect in the naturally infected consistently exceeded power at a population-level and increased more rapidly with trial size. At a trial size of 20,000, power was 11% for the naturally infected effect compared to 6% for the population effect; at trial size 80,000, power increased to 25% for the naturally infected but only to 7% at the population-level (Figure 3). Trial sizes needed to detect growth effects were considerably larger than those required to achieve 90% power for the primary endpoint, which ranged from 919 to 5,642 across all simulation settings.

**Figure 3:**
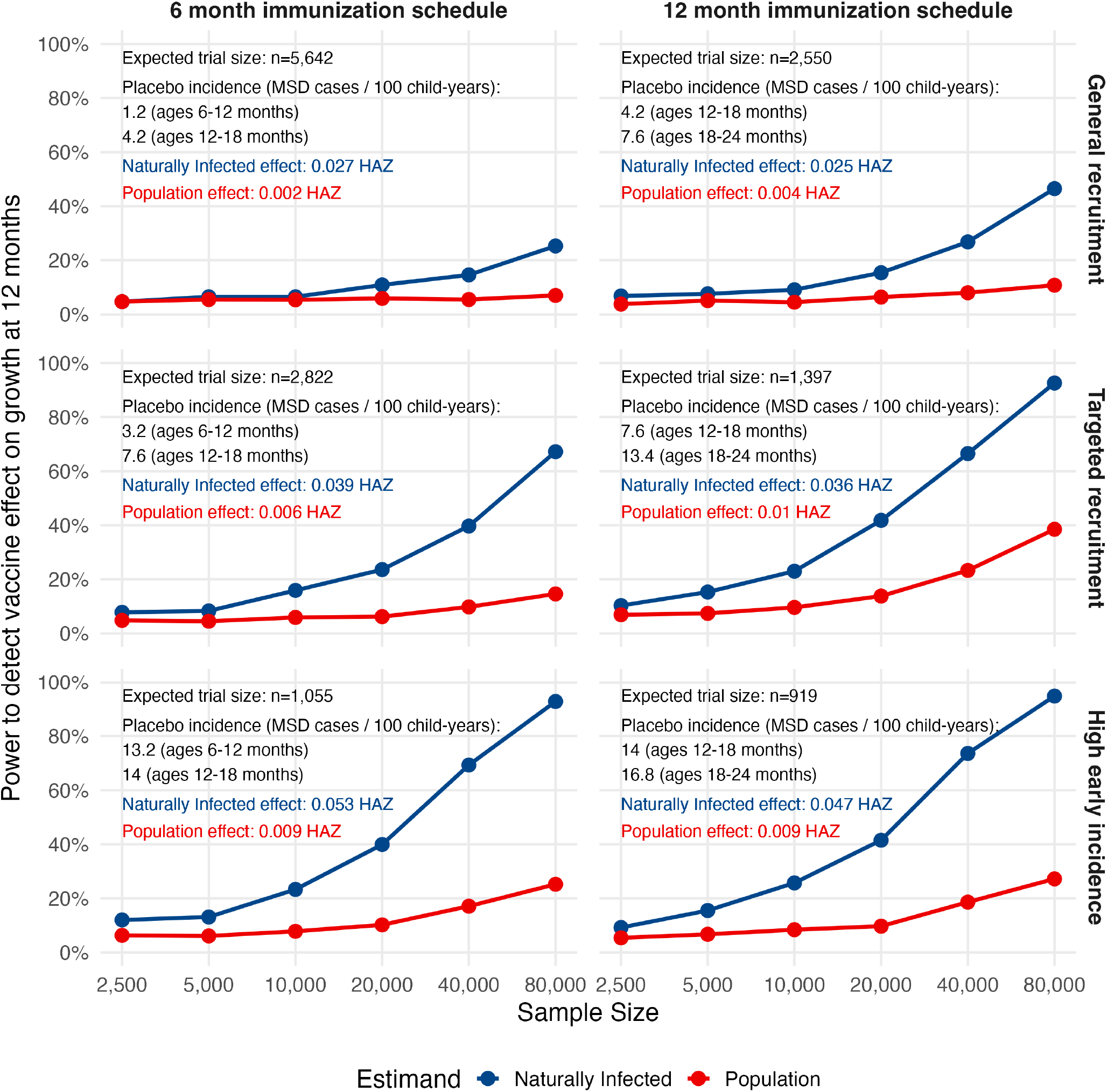
Power to detect a vaccine effect on HAZ at the end of a *Shigella* vaccine trial comparing 6-month versus 12-month immunization schedules across three recruitment strategies. Expected trial size denotes the sample size required to achieve 90% power for the primary endpoint of moderate-to-severe *Shigella*-attributable diarrhea under each trial configuration. Reported incidence corresponds to moderate-to-severe *Shigella*-attributable diarrhea in the first and second halves of follow-up, defined by participants’ ages during each period. Estimates are shown for both naturally infected (blue) and population-level (red) effects of *Shigella* vaccination on HAZ.

The population-level estimates were also more likely to show a negative effect of vaccination on growth. At n=20,000, 43% of population-level estimates were negative compared with 26% of naturally infected estimates (Figure 4). 1.9% of population-level estimates were statistically significant in the negative direction, while 0.4% of the naturally infected estimates were significant (Figure S15).

**Figure 4:**
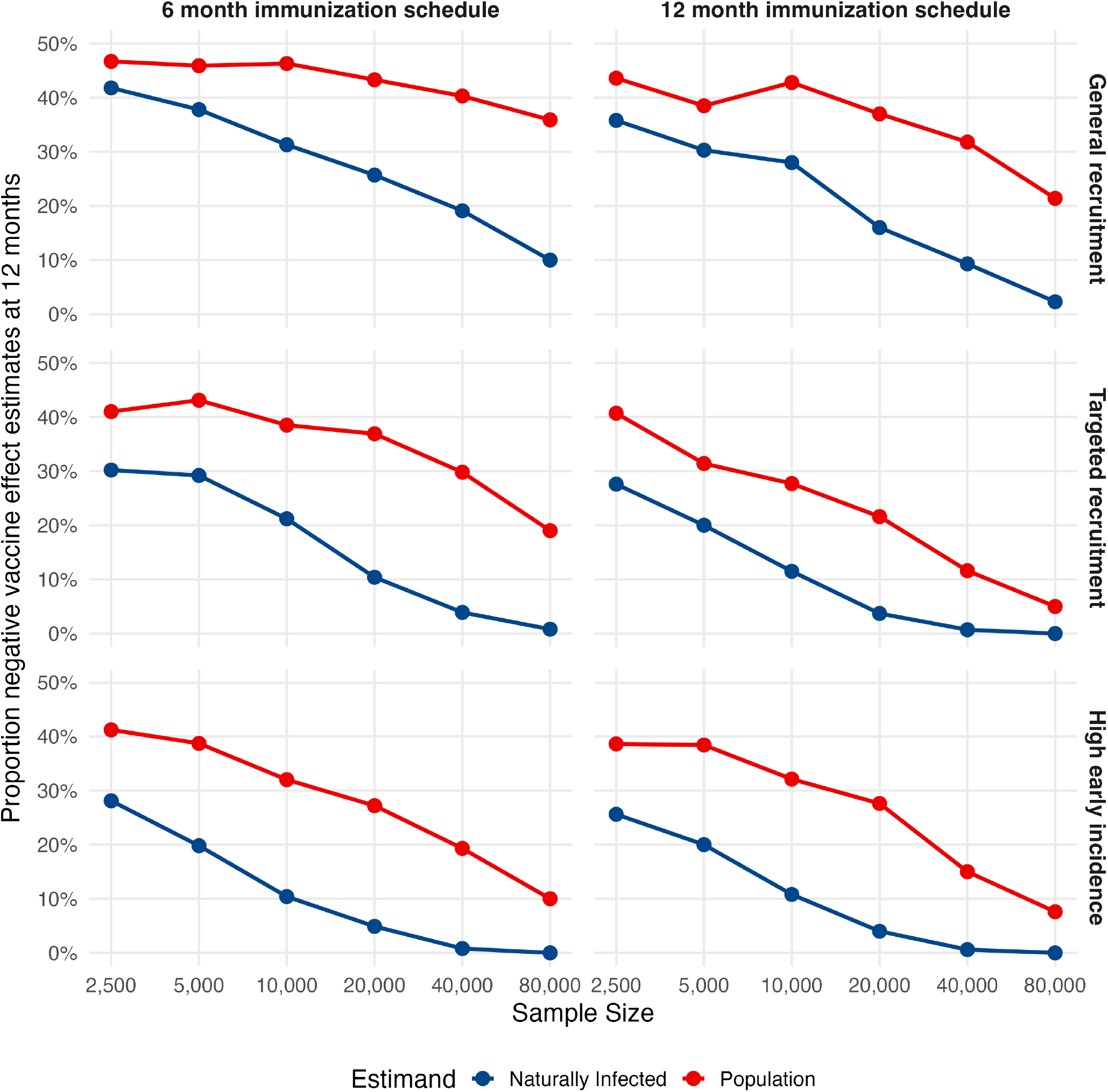
Proportion of negative point estimates in trials using 6 month vs 12 month immunization schedules under three different settings. Naturally infected estimates are shown in blue, while population-level estimates are shown in red.

### 2.2 Trial Design

#### 2.2.1 Immunization schedule

The effect size in the naturally infected was slightly larger under 6 month immunization schedules than 12 month immunization schedules in all simulation settings. However, trials using a 12 month immunization schedules were consistently better powered and returned fewer negative point estimates.

In the targeted recruitment scenario, power to detect a vaccine effect at 12 months with a sample size of 20,000 was 24% under 6 month immunization schedules and 42% under 12 month immunization schedules 3). Estimates were negative in 10% of simulations under 6 month schedules, compared with 4% under 12 month schedules. At the largest trial size of 80,000, power increased to 67% and 93%, respectively.

The difference between immunization schedules was less apparent in the scenario with high early incidence; power and proportion of negative point estimates for the 6 month immunization schedules were comparable to that for the 12 month immunization schedules. At a trial size of 20,000, power was 40% under the 6-month schedule and 42% under the 12-month schedule, with 5% and 4% negative estimates, respectively (Figures 3, 4).

#### 2.2.2 Recruitment strategy

Targeted recruitment strategies improved power over general recruitment. Under 12 month immunization schedules, targeted recruitment led to 67% power to detect a naturally infected vaccine effect at 12 months with trial size 40,000 compared to 27% under the general recruitment strategy (3). Correspondingly, the general recruitment strategy had a higher proportion of negative point estimates (9%) than the targeted recruitment setting (1%) (Figure 4). Similarly, there was higher power and lower risk of negative point estimates in the high early incidence setting compared to the general recruitment strategy.

#### 2.2.3 Interim measurements

Incorporating interim measurements did not improve power over a single outcome measurement at month 12. In the targeted recruitment setting, power for the 6 month outcome alone was much lower than the month 12 outcome alone (10% at 6 months vs 42% at 12 months at n=20,000 for the 12 month vaccine schedule; Figure 5). Averaging effects across both time points (29% power) did not improve power over the 12 month outcome alone.

**Figure 5:**
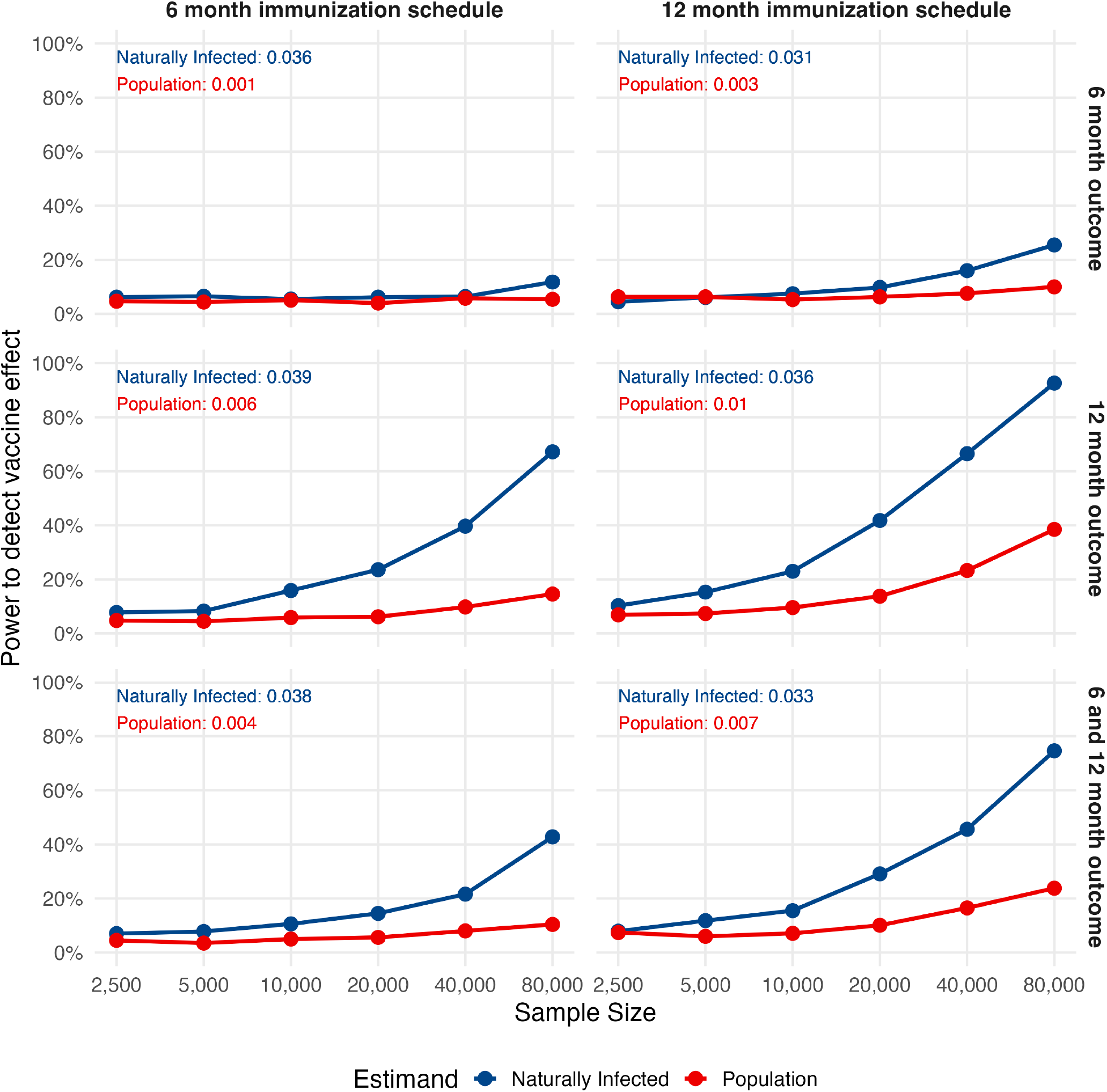
Power for growth effects at 6 months of follow-up, 12 months of follow-up, and when incorporating both measurements at 6 and 12 months under a targeted recruitment schedule for both 6 month and 12 month immunization schedules. Naturally infected estimates are shown in blue, while population-level estimates are shown in red.

## Discussion

Estimation of the effect of *Shigella* vaccination on linear growth is likely to be underpowered at realistic Phase 3 trial sizes based on power calculations for the primary endpoint of moderate-to-severe diarrhea. Even in the most favorable scenarios using the naturally infected approach, we estimate the required sample size to achieve 80% power for a vaccine effect on growth to be around 80,000, which is likely not achievable in a Phase 3 trial setting. Low power for growth effects at realistic trial sizes poses a risk of estimating null or even significantly negative effects, complicating the communication of vaccine benefit in an already sensitive vaccine policy landscape. These findings suggest that post-marketing studies will be necessary for evaluating the effectiveness of *Shigella* vaccines on linear growth.

Estimating the effect of *Shigella* vaccination on linear growth in the naturally infected population increases effect size, improves power, and decreases risk of negative estimates compared to a traditional population-level effect. As expected, population-level effect sizes were five to ten times smaller than the naturally infected effect sizes, reflecting dilution from children who did not have *Shigella* diarrhea during follow-up and therefore were not at risk of *Shigella*-attributable growth faltering. This dilution led to a high proportion of negative point estimates, with a non-trivial fraction reaching statistical significance despite a small positive true effect. These findings emphasize the risk that population-level analyses could falsely suggest harm of the vaccine on growth. In contrast, the naturally infected approach mitigates these risks while more directly capturing the benefit of vaccination among those at risk of *Shigella*-attributable growth-faltering.

Beyond the estimation approach, trial designs that target high risk children can further maximize likelihood of detecting an effect of vaccination on growth. This may include structuring enrollment criteria to prioritize groups with higher incidence and larger anticipated effects of *Shigella* diarrhea on growth. Choice of immunization schedules is more nuanced; while the true effect size of *Shigella* vaccination on growth outcomes among the naturally infected was slightly larger under earlier immunization schedules, power to detect this effect was generally higher under later immunization schedules. This pattern is likely driven by lower incidences at younger ages in most sites, resulting in fewer cases observed in a clinical trial setting with earlier follow-up (i.e., a smaller naturally infected subgroup). In settings like our simulated high early incidence scenario, differences between immunization schedules were less pronounced. This finding suggests that if earlier immunization schedules are planned, targeting sites with high incidence early in life will be critical to ensure a sufficiently large naturally infected subgroup.

Incorporating interim growth measurements did not increase effect size or improve power to detect an effect of *Shigella* vaccination on growth. While non-intuitive, this may be driven by the longer duration of follow-up needed to observe the peak effect of infection on growth for severe cases (Figure 2), as well as correlation of growth measurements over time, which limits additional information contributed by interim assessments. As a result, collection of interim growth measurements is unlikely to be worth the allocation of resources and may even make it more difficult to show a vaccine effect on growth.

Our analysis is limited by the uncertainty surrounding immunization schedule, vaccine efficacy, and serotype coverage for *Shigella* vaccines. Our simulations assume a quadrivalent vaccine formulation incorporated in several existing candidates.^8, 19, 20^ Evaluation of a bivalent vaccine would be expected to have lower power due to lower incidence of disease caused by vaccine-covered serotypes. Additionally, while we defined MSD in accordance with the GEMS definition, alternative definitions based on the modified Vesikari score and/or dysentery are under consideration.^17^ We expect power to be comparable for trials with alternative severity definitions given the diarrhea incidence estimates are similar across definitions.^18^ To facilitate evaluation of alternative scenarios, all simulation code is publicly available at https://github.com/allicodi/shigella_vax_growth_sim.

While our findings suggest that clinical trials aiming to detect an effect of *Shigella* vaccination on linear growth will almost certainly be underpowered, careful selection of estimation approach and trial design can reduce the risk of null or misleading results and better capture the true value of the vaccine. These results should inform both the design and analysis of future Phase 3 trials, with the expectation that post-licensure studies will ultimately be required for well powered estimates of vaccine effects on growth.

## Data Availability

MAL-ED data are available online through ClinEpiDB (https://clinepidb.org/ce/app/workspace/analyses/DS_5c41b87221/new). EFGH data are available online in Vilvi (https://doi.org/10.25934/PR00011860).

https://doi.org/10.25934/PR00011860

https://github.com/allicodi/shigella_vax_growth_sim

## Research in context

### Evidence before this study

We searched PubMed on March 31, 2026 for articles published in any language since January 1, 1990 using the search terms (shigell*) AND (vaccine) AND (grow* OR linear OR height OR length OR stunt*) AND (pediatric OR paediatric OR infant* OR child*) AND (“phase 3” OR “phase III” OR trial OR randomized). We identified 25 publications, none of which evaluated methods or projected estimates of the impact of *Shigella* vaccines on linear growth faltering in the context of a Phase 3 vaccine trial.

### Added value of this study

We describe statistical methods that increase power for detecting a positive vaccine growth effect in a clinical trial. Using a realistic simulation study parameterized by recent large observational studies of diarrheal disease, we also illustrate the impact of trial recruitment strategy and other design features that may impact power to detect vaccine effects on growth. Our results support decision making regarding vaccine scheduling, recruitment strategies, and statistical analysis approaches for upcoming Phase 3 trials of *Shigella* vaccines.

### Implications of all the available evidence

Our simulations demonstrate that power to detect positive growth effects is likely to be limited in realistic trial sizes. Power can be improved by using targeted statistical methods and recruiting children at high risk for growth faltering; however, even in the most optimistic settings, the trial size required to be well powered for a linear growth endpoint is well beyond expected sizes of Phase 3 trials. As such, investigators should carefully weigh the risks and benefits of including linear growth measurements as a secondary endpoint in a clinical trial.

### Contributors

AC contributed to the curation of data, formal analysis, methodology development, software development, visualization, writing the initial draft of the manuscript. DB and LRM contributed to the conceptualization, methodology development, project administration, funding acquisition, validation, and reviewing/editing the manuscript.

