## Supplemental Material for "Estimating the impact of *Shigella* vaccines on growth outcomes and implications for clinical trial design"

### 1 Statistical Methods

#### 1.1 Principal Stratification

Comparing post-infection outcomes between vaccinated and unvaccinated individuals can be difficult. A naive comparison of post-infection outcomes between individuals infected in each trial arm is generally not an appropriate causal effect due to the potential for systematic differences that induce selection bias. For example, participants who are infected in the vaccine arm may have weaker immune systems on average than infected participants in the control arm, leading to worse post-infection outcomes on average (1). Similarly, infections that occur in the vaccine arm may disproportionately involve serotypes not targeted by the vaccine, such that differences in post-infection outcome may be mediated through the effects of different serotypes (2).

We use an approach known as *principal stratification* (3) to estimate a causal effect of *Shigella* vaccine on the post-infection outcome of linear growth. This approach allows us define subgroups termed *principal strata* based an individual's potential infection status under both vaccine and placebo. We can define four principal strata based on combinations of infection status under vaccine and placebo:

1. *Immune* - individuals who are never infected
2. *Protected* - individuals who are infected under placebo, but not under vaccine
3. *Harmed* - individuals individuals who are not infected under placebo, but are infected under vaccine
4. *Doomed* - individuals who are always infected

In a Phase 3 clinical trial, it is reasonable to assume that the vaccine will not cause any individuals to have an infection, i.e. no participants fall into the *harmed* stratum. Therefore, we are left with three principal strata. We can condition on these strata to estimate effects without inducing selection bias, as we treat them as pre-specified subgroups.

Previous vaccine studies have applied principal stratification to estimate effects of vaccine on post-infection outcomes in the *doomed* principal stratum (2; 4; 5). Restricting to this stratum is appropriate for endpoints that are only defined for individuals with an infection, such as set point viral load. However, some post-infection outcomes of interest, such as linear growth, are defined regardless of infection. While vaccination may still benefit individuals in the doomed stratum by decreasing severity of infection thereby reducing growth faltering, restricting analyses to this subgroup would omit participants whose infection is prevented by the vaccine, i.e. those who would be at risk of *Shigella*-attributable growth faltering under placebo but not under vaccine. Excluding these participants misses perhaps the largest component of the vaccine effect. Conversely, a population-level comparison including all three strata would dilute the observed effect, as immune individuals are unlikely to have their outcome affected by vaccination assuming the only way the vaccine can improve growth is through effects on infection. We therefore focus on the *naturally infected* subset–a combination of the doomed and protected strata. This subset captures the effect of vaccination on growth among children who would be infected in absence of vaccine.

Figure S1 is an extension on Figure 1 from the main text detailing the principal strata for each child in a hypothetical study. In this example, the magnitude of the naturally infected vaccine effect is larger than both the doomed and population-level effects.

#### 1.2 Identification Assumptions

As we can only observe each participant under the treatment they are randomized to, we rely on several assumptions to identify the subset of children who would be infected in absence of vaccination, i.e. the *naturally infected* subgroup.

The following assumptions are required. These assumptions are common to many studies of infectious diseases in randomized trials.

1. *No interference*: One participant's vaccination status does not affect another participant's risk of infection nor growth outcomes. In a Phase 3 trial, participants typically represent a small fraction of the broader population, and the vaccine is not available outside the study, making this assumption plausible.

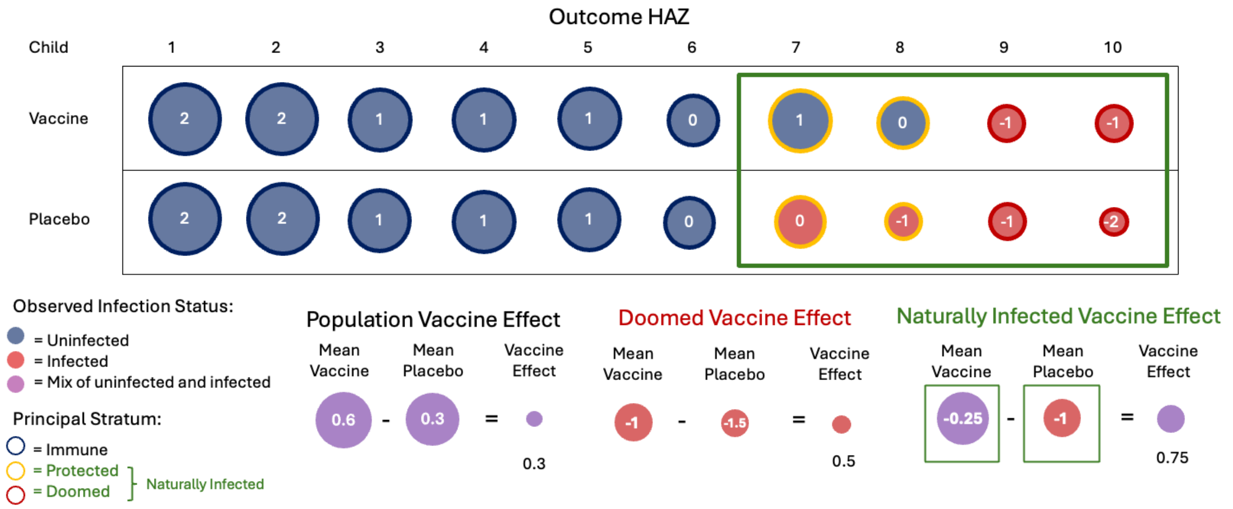

Figure 1: **Illustration of population-level and naturally infected estimands broken into principal strata.** Each circle represents a child's growth outcome under vaccine or placebo. Blue circles denote children who remain uninfected, and red circles denote children who experience *Shigella*-attributable diarrhea. The outlines of the circles indicate which principal stratum each child belongs to–doomed (red), protected (yellow), immune (blue). The green box identifies children who would be infected under placebo, corresponding to the naturally infected subset.

2. *Causal consistency*: The outcomes observed under an assigned treatment (placebo or vaccine) correspond to the outcomes that would be experienced under that same treatment, provided it is applied in a well-defined and consistent manner. In a Phase 3 trial, procedures for administering the vaccine and placebo are protocolized and typically consistency of vaccine manufacturing processes has been assured.

3. *Monotonicity*: Vaccination does not increase the risk of infection. That is, individuals who would not have been infected under placebo also would not be infected under vaccine. This is a reasonable assumption for vaccines that have advanced to large-scale trials and eliminates the harmed stratum.

4. *Randomization*: Treatment assignment is independent of participants' underlying characteristics and potential outcomes. This is ensured by the randomized design of the trial.

5. *Positivity for infection under placebo*: There is a non-zero chance of each participant being infected under placebo. This is ensured by conducting the trial in a site with ongoing transmission.

Additionally, we require at least one of the following assumptions:

1. *Exclusion Restriction*: There is no effect of vaccination on growth outcomes in absence of infection. This assumption implies that any effect of the vaccine on growth operates through preventing or modifying infection. We think this is plausible as participants without *Shigella* diarrhea are not expected to experience *Shigella*-attributable growth faltering. However, this may be violated if the vaccine prevents subclinical infections that cause faltering, or provides cross protection against other pathogens that influence growth.

2. *Partial Principal Ignorability*: After accounting for measured baseline characteristics, there are no unmeasured common causes of both infection and growth faltering. While this assumption is strong in practice, it may be possible to minimize bias resulting from violations of this assumption through deliberate collection of baseline covariate information from trial participants.

#### 2 Additional parameterization details

##### 2.1 Baseline HAZ

| | Immunization schedule | |
|---|---|---|
| **Setting** | **6 month** | **12 month** |
| General & targeted recruitment (The Gambia) | -0.56 (1.14) | -0.94 (1) |
| High early incidence (Peru) | -0.71 (1.13) | -0.78 (0.97) |

Table 1: **Baseline HAZ in each simulation setting by immunization schedule.** Mean baseline HAZ (SD)

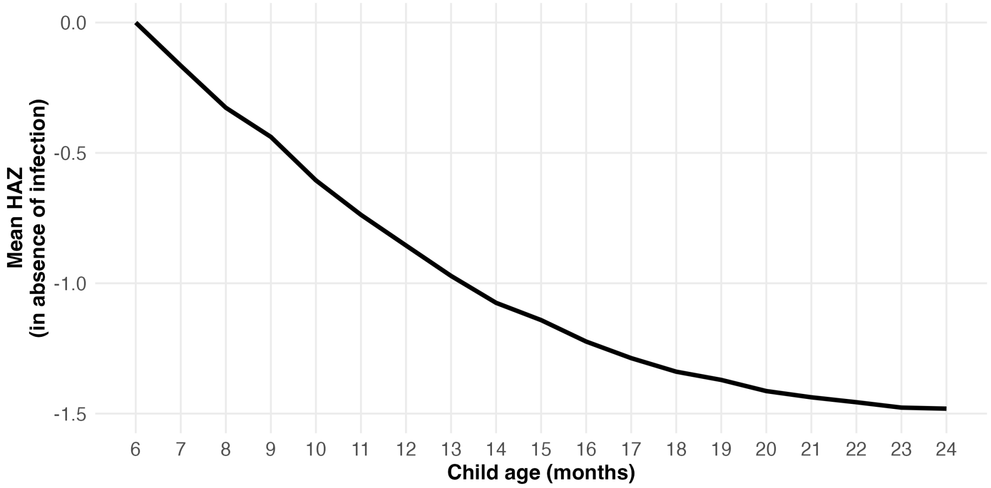

Figure 2: **Average conditional growth trajectory in absence of infection used in all simulation settings**. Shown for child with baseline HAZ of 0

##### 2.2 Incidence

| | 6–12 months | | 12–18 months | | 18–24 months | |
|---|---|---|---|---|---|---|
| **Setting** | **MAD** | **MSD** | **MAD** | **MSD** | **MAD** | **MSD** |
| General recruitment | 4.2 | 1.2 | 13.2 | 4.2 | 24.8 | 7.6 |
| Targeted recruitment | 8.6 | 3.2 | 22.4 | 7.6 | 40.4 | 13.4 |
| High early incidence | 20.0 | 13.2 | 15.4 | 14.0 | 22.8 | 16.8 |

Table 2: ***Shigella* diarrhea incidence per 100 child-years in each simulation setting by age group**

#### 2.3 Incidence rate ratio by baseline HAZ

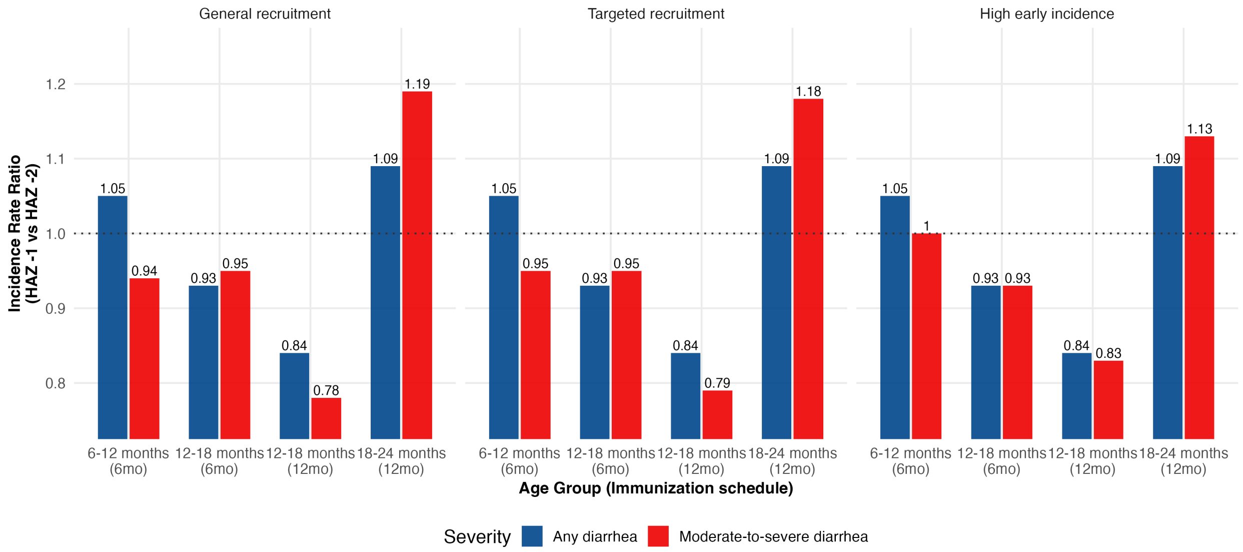

Figure 3: ***Shigella* incidence rate ratios by baseline HAZ.** Medically attended *Shigella* diarrhea (blue) and moderate-to-severe *Shigella* diarrhea (red) for a one unit increase of HAZ from -2 to -1 by age group and immunization schedule in each simulation setting

#### 2.4 Study design specific parameterizations

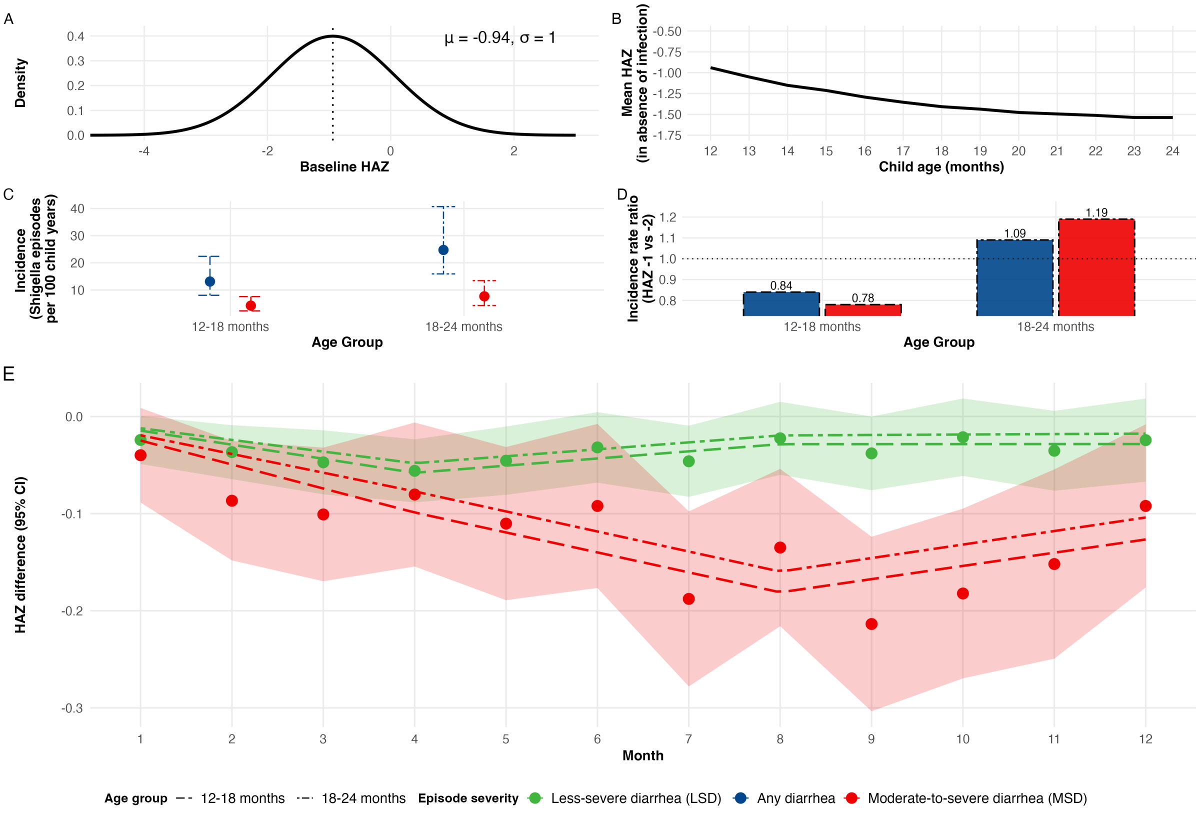

Figure 4: **Simulation parameterization for general recruitment strategy, 12 month immunization schedule**. (A) Distribution of baseline HAZ. (B) One-year average growth trajectory in absence of infection for children with mean baseline HAZ. (C) Average incidence of MAD and MSD in young children (first 6 months of follow up for a trial enrolling 12 month olds) and older children (second half of the trial). (D) Incidence was simulated to vary by HAZ. The incidence rate ratio for a one-unit increase of baseline HAZ from -2 to -1 for any *Shigella* diarrhea by age group is shown. (E) Monthly estimated effects of *Shigella* diarrhea by severity level (dash, dot-dash). Splines were fit for each age strata separately, with younger children experiencing larger deleterious growth effects following *Shigella* diarrhea.

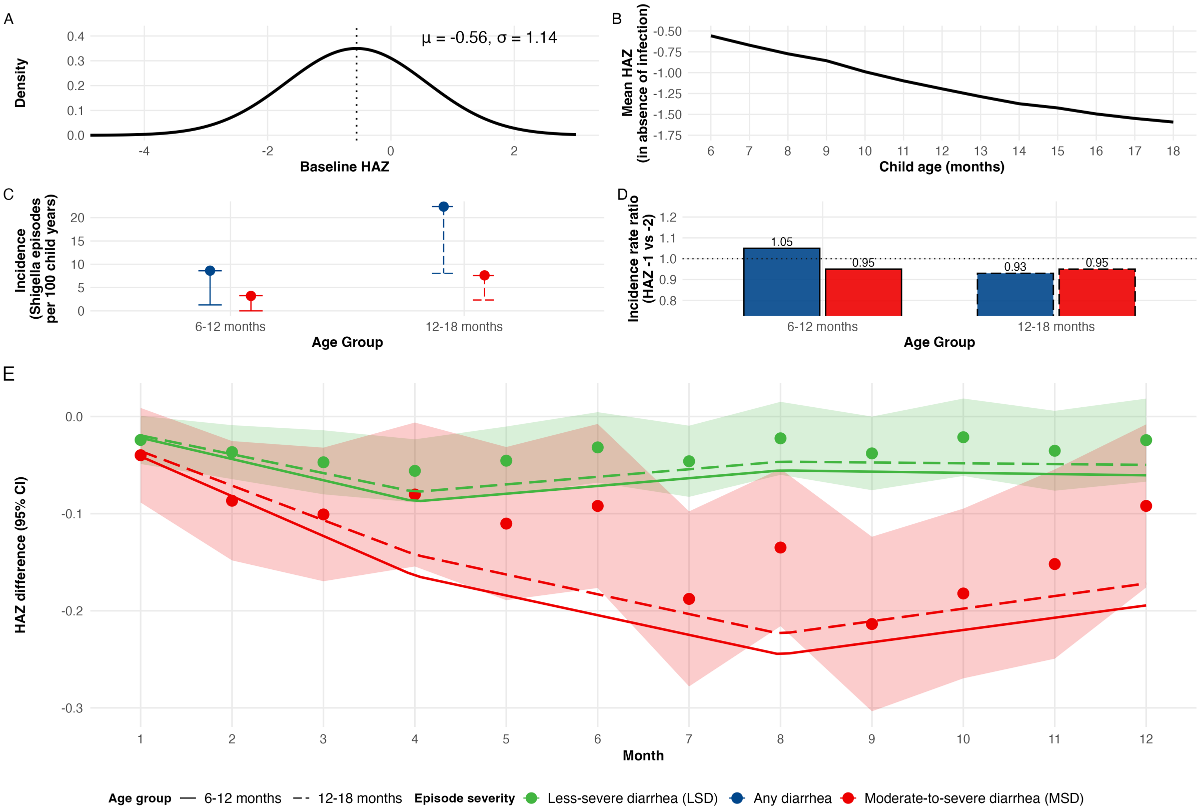

Figure 5: **Simulation parameterization for targeted recruitment strategy, 6 month immunization schedule**. (A) Distribution of baseline HAZ. (B) One-year average growth trajectory in absence of infection for children with mean baseline HAZ. (C) Average incidence of MAD and MSD in young children (first 6 months of follow up for a trial enrolling 6 month olds) and older children (second half of the trial). (D) Incidence was simulated to vary by HAZ. The incidence rate ratio for a one-unit increase of baseline HAZ from -2 to -1 for any *Shigella* diarrhea by age group is shown. (E) Monthly estimated effects of *Shigella* diarrhea by severity level (bold, dash). Splines were fit for each age strata separately, with younger children experiencing larger deleterious growth effects following *Shigella* diarrhea.

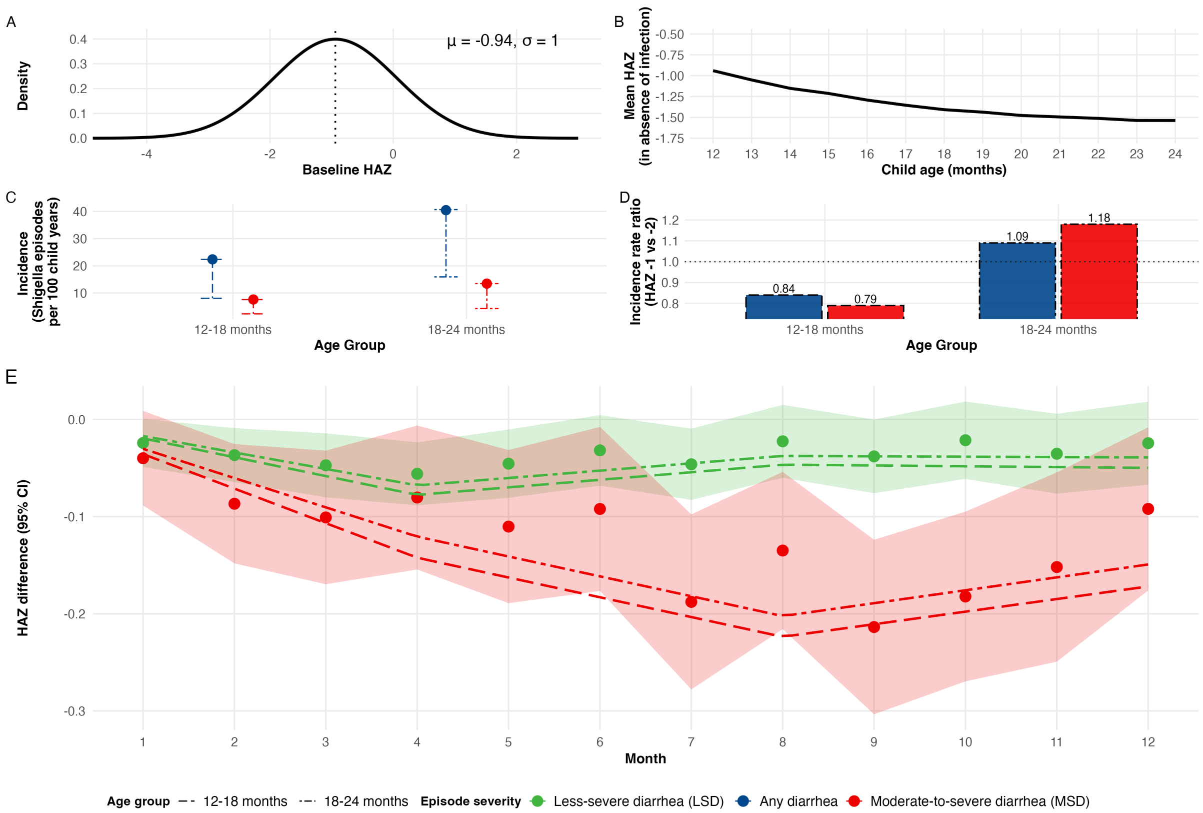

Figure 6: **Simulation parameterization for targeted recruitment strategy, 12 month immunization schedule**. (A) Distribution of baseline HAZ. (B) One-year average growth trajectory in absence of infection for children with mean baseline HAZ. (C) Average incidence of MAD and MSD in young children (first 6 months of follow up for a trial enrolling 12 month olds) and older children (second half of the trial). (D) Incidence was simulated to vary by HAZ. The incidence rate ratio for a one-unit increase of baseline HAZ from -2 to -1 for any *Shigella* diarrhea by age group is shown. (E) Monthly estimated effects of *Shigella* diarrhea by severity level (dash, dot-dash). Splines were fit for each age strata separately, with younger children experiencing larger deleterious growth effects following *Shigella* diarrhea.

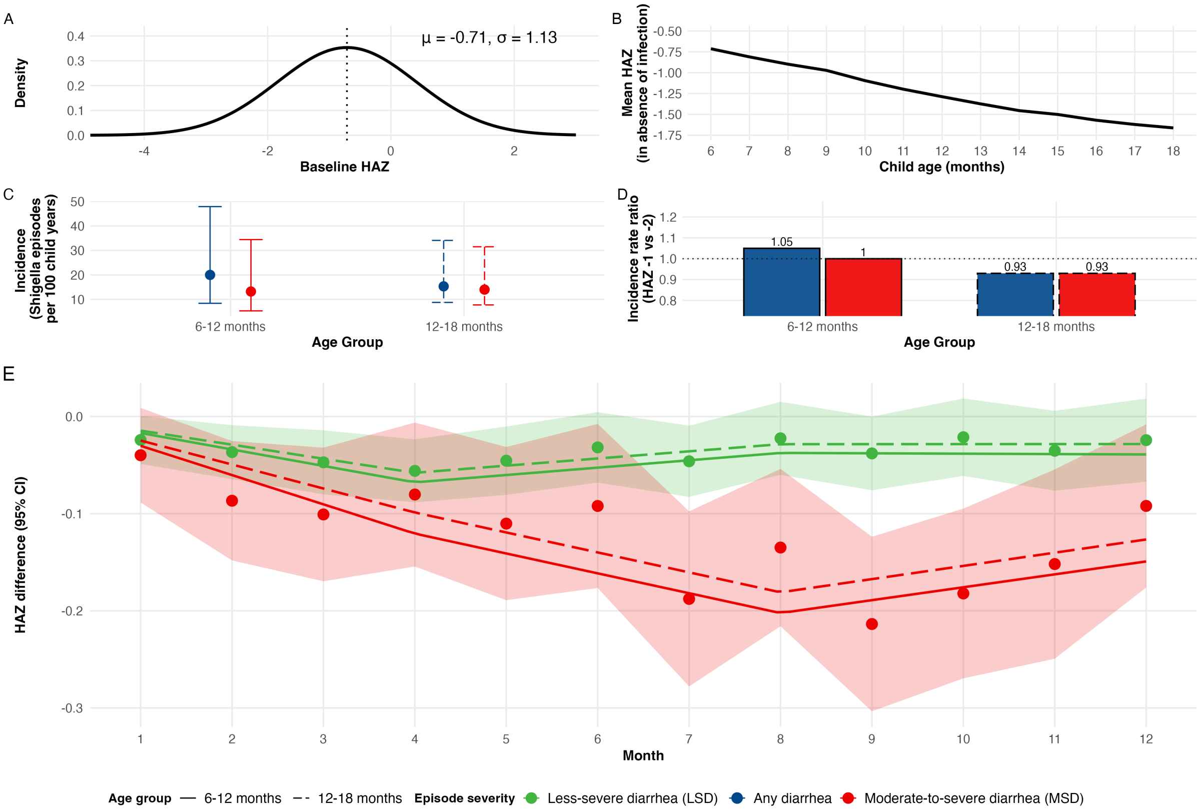

Figure 7: **Simulation parameterization for high early incidence strategy, 6 month immunization schedule**. (A) Distribution of baseline HAZ. (B) One-year average growth trajectory in absence of infection for children with mean baseline HAZ. (C) Average incidence of MAD and MSD in young children (first 6 months of follow up for a trial enrolling 6 month olds) and older children (second half of the trial). (D) Incidence was simulated to vary by HAZ. The incidence rate ratio for a one-unit increase of baseline HAZ from -2 to -1 for any *Shigella* diarrhea by age group is shown. (E) Monthly estimated effects of *Shigella* diarrhea by severity level (bold, dash). Splines were fit for each age strata separately, with younger children experiencing larger deleterious growth effects following *Shigella* diarrhea.

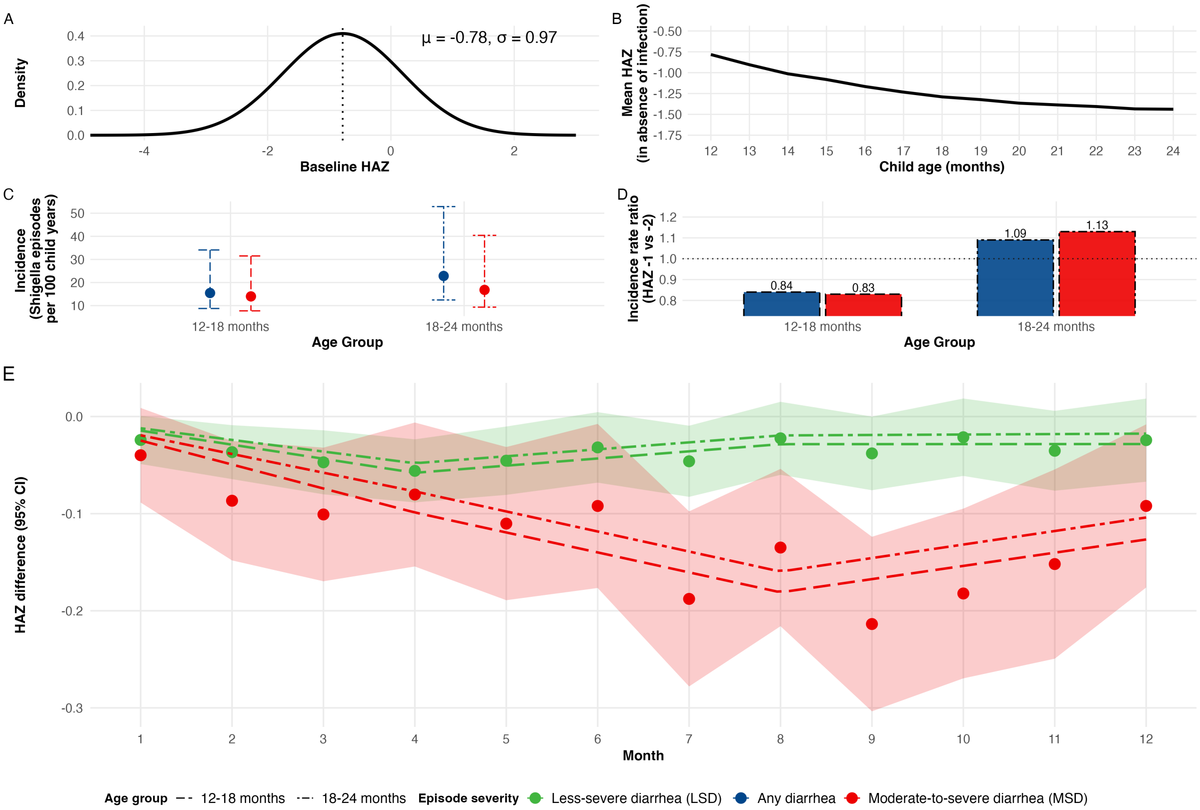

Figure 8: **Simulation parameterization for high early incidence strategy, 12 month immunization schedule**. (A) Distribution of baseline HAZ. (B) One-year average growth trajectory in absence of infection for children with mean baseline HAZ. (C) Average incidence of MAD and MSD in young children (first 6 months of follow up for a trial enrolling 12 month olds) and older children (second half of the trial). (D) Incidence was simulated to vary by HAZ. The incidence rate ratio for a one-unit increase of baseline HAZ from -2 to -1 for any *Shigella* diarrhea by age group is shown. (E) Monthly estimated effects of *Shigella* diarrhea by severity level (dash, dot-dash). Splines were fit for each age strata separately, with younger children experiencing larger deleterious growth effects following *Shigella* diarrhea.

#### 3 Results

##### 3.1 True effect size

| | Outcome | General recruitment | | Targeted recruitment | | High early incidence | |
|---|---|---|---|---|---|---|---|
| | | 6 mo | 12 mo | 6 mo | 12 mo | 6 mo | 12 mo |
| Naturally Infected | $Y_3$ | 0.014 | 0.012 | 0.020 | 0.017 | 0.018 | 0.017 |
| | $Y_6$ | 0.025 | 0.022 | 0.036 | 0.031 | 0.035 | 0.035 |
| | $Y_9$ | 0.023 | 0.023 | 0.035 | 0.033 | 0.048 | 0.044 |
| | $Y_{12}$ | 0.027 | 0.025 | 0.039 | 0.036 | 0.053 | 0.047 |
| | $Y_{6–12}$ | 0.026 | 0.023 | 0.038 | 0.033 | 0.044 | 0.041 |
| | $Y_{3–6–9–12}$ | 0.022 | 0.020 | 0.032 | 0.029 | 0.039 | 0.036 |
| Population | $Y_3$ | <0.001 | <0.001 | <0.001 | <0.001 | <0.001 | <0.001 |
| | $Y_6$ | <0.001 | 0.001 | 0.001 | 0.003 | 0.003 | 0.003 |
| | $Y_9$ | 0.001 | 0.003 | 0.003 | 0.007 | 0.006 | 0.006 |
| | $Y_{12}$ | 0.002 | 0.004 | 0.006 | 0.010 | 0.009 | 0.009 |
| | $Y_{6–12}$ | 0.001 | 0.003 | 0.004 | 0.007 | 0.006 | 0.006 |
| | $Y_{3–6–9–12}$ | 0.001 | 0.002 | 0.003 | 0.005 | 0.005 | 0.004 |

Table 3: **Effect of *Shigella* vaccination on height-for-age z-score under each simulation setting.** Shown are the effects at each outcome time of interest, where $Y_m$ denotes height-for-age z-score measured $m$ months after infection. Outcomes with more than one $m$ averaged effects across multiple outcome time points. Columns correspond to each simulation setting under immunization schedules with full protection at 6 or 12 months of age.

##### 3.2 Power

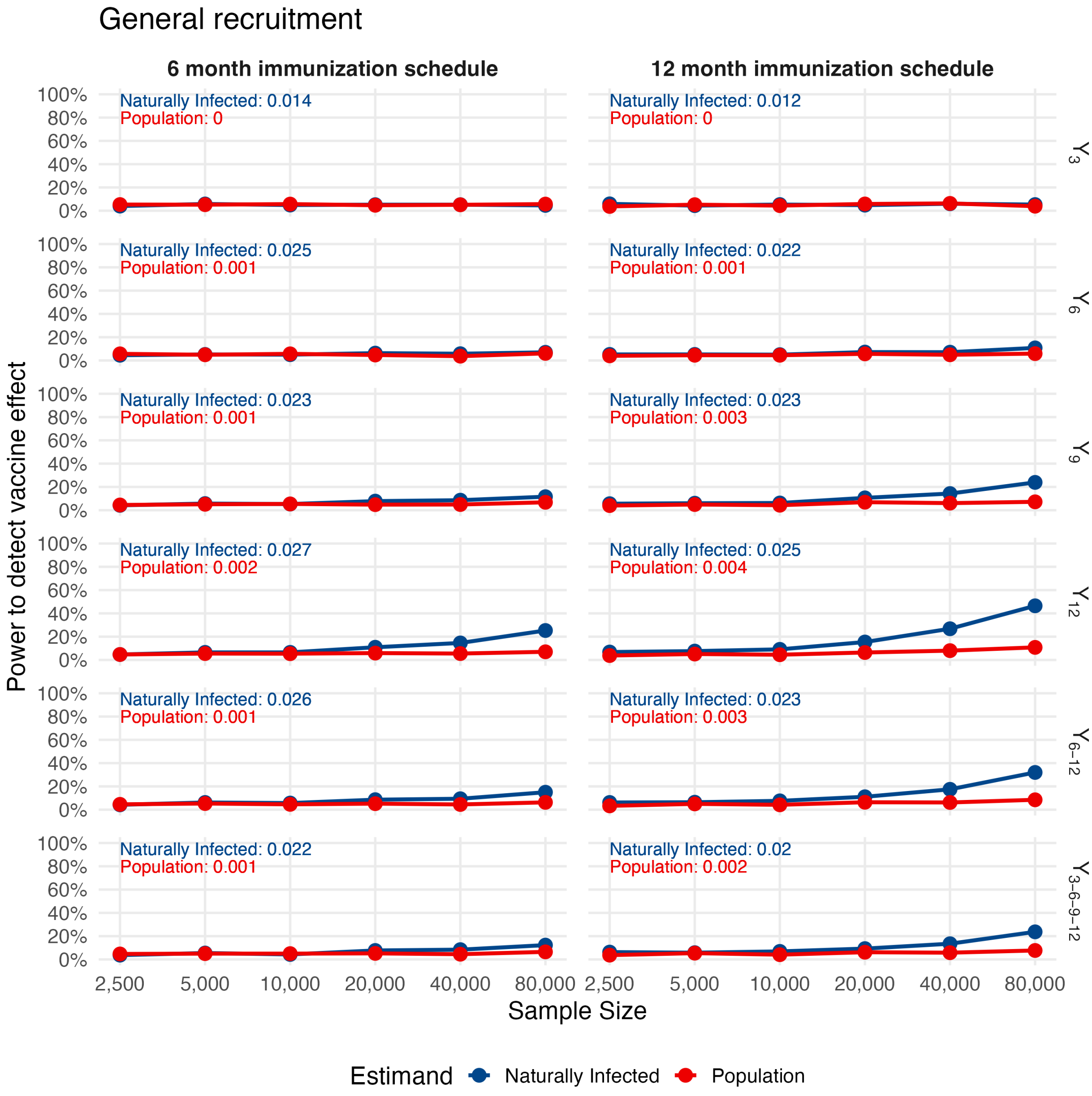

Figure 9: **Power for all endpoints under general recruitment strategy.** Shown are power estimates for the effects at each outcome time of interest, where $Y_m$ denotes height-for-age z-score measured $m$ months after infection. Columns correspond to immunization schedules with full protection at 6 or 12 months of age. Naturally infected estimates are shown in blue, while population-level estimates are shown in red.

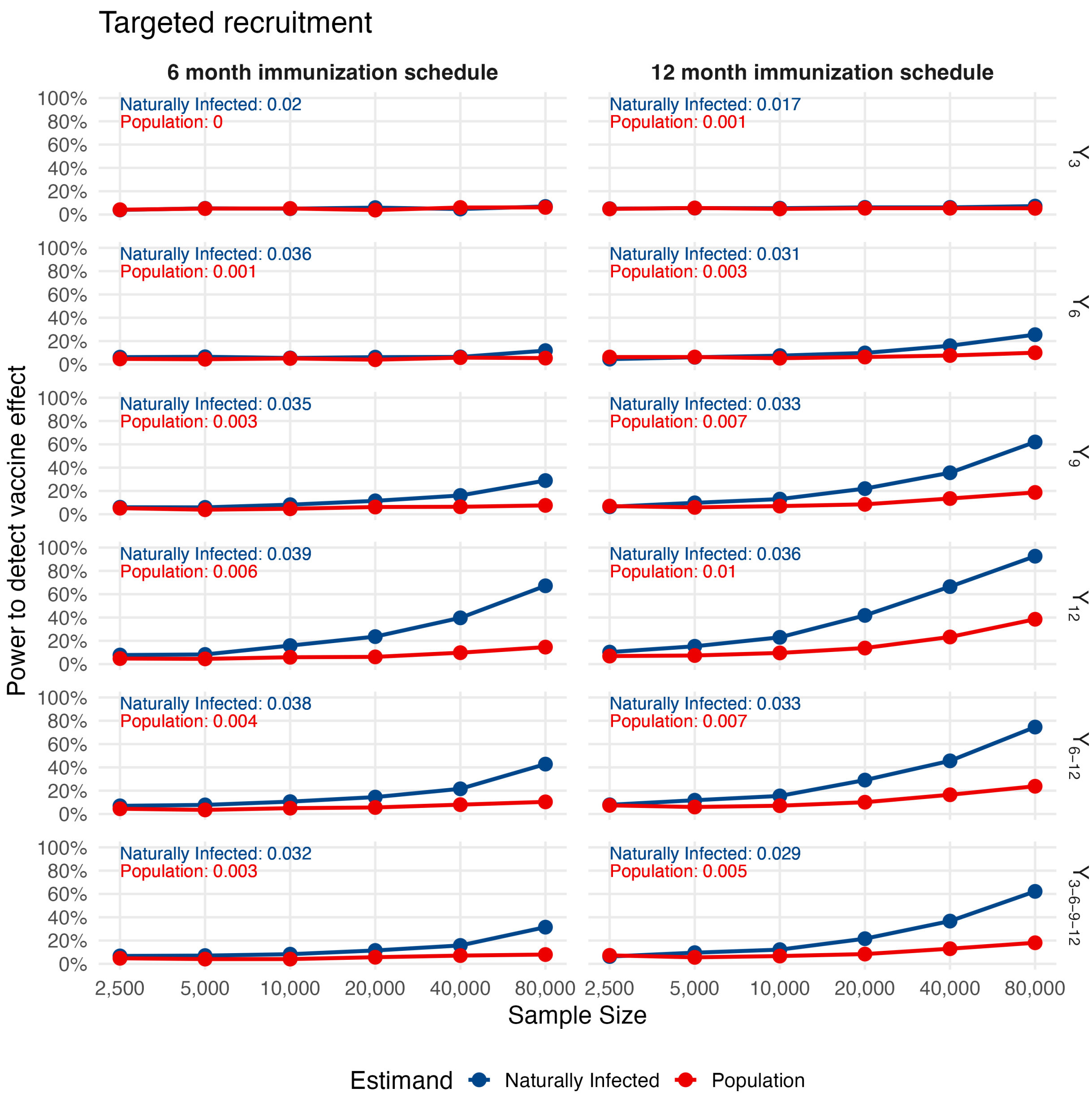

Figure 10: **Power for all endpoints under targeted recruitment strategy.** Shown are power estimates for the effects at each outcome time of interest, where $Y_m$ denotes height-for-age z-score measured $m$ months after infection. Columns correspond to immunization schedules with full protection at 6 or 12 months of age. Naturally infected estimates are shown in blue, while population-level estimates are shown in red.

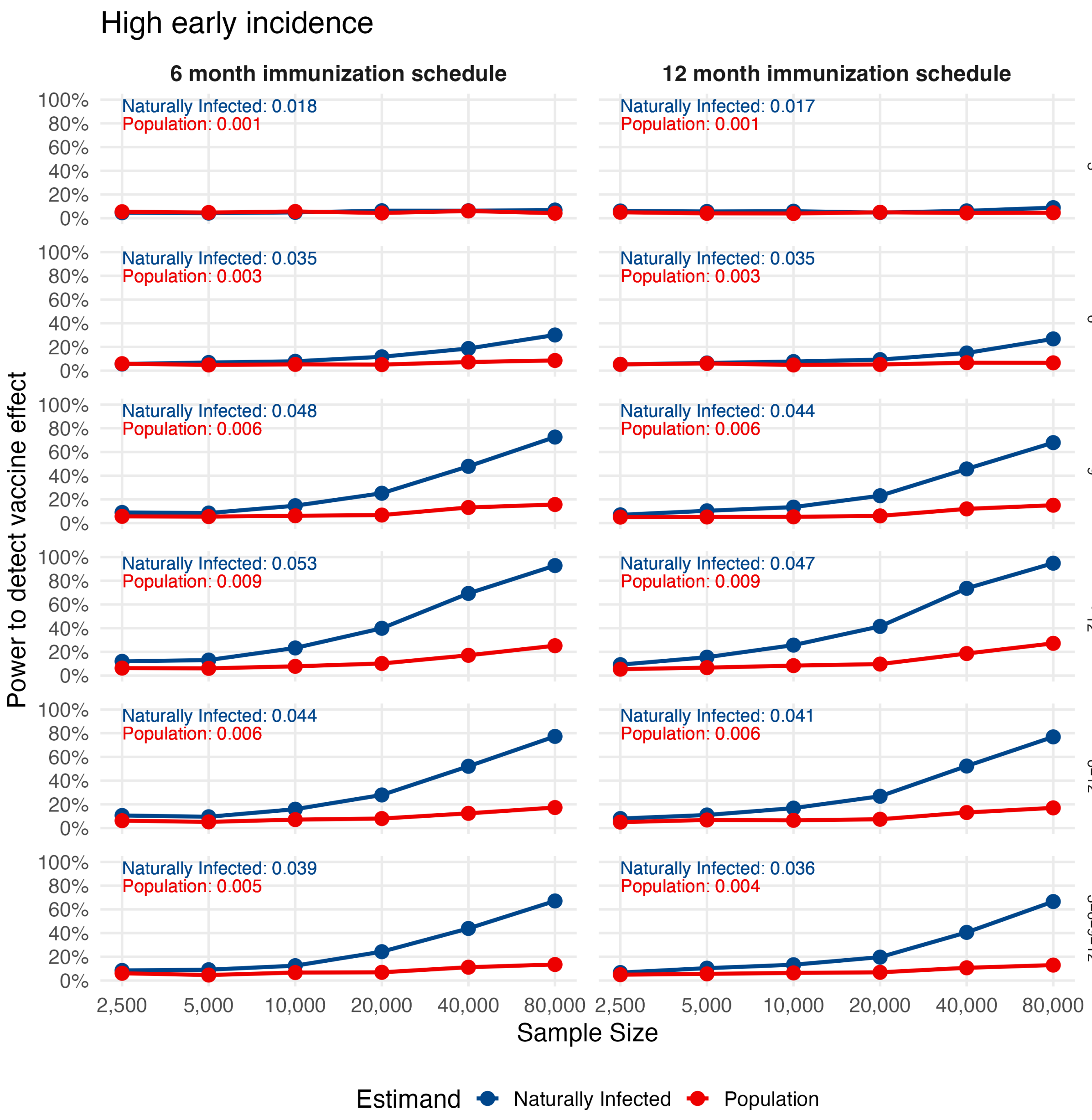

Figure 11: **Power for all endpoints under high early incidence recruitment strategy.** Shown are power estimates for the effects at each outcome time of interest, where $Y_m$ denotes height-for-age z-score measured $m$ months after infection. Columns correspond to immunization schedules with full protection at 6 or 12 months of age. Naturally infected estimates are shown in blue, while population-level estimates are shown in red.

##### 3.3 Proportion of negative estimates across simulations

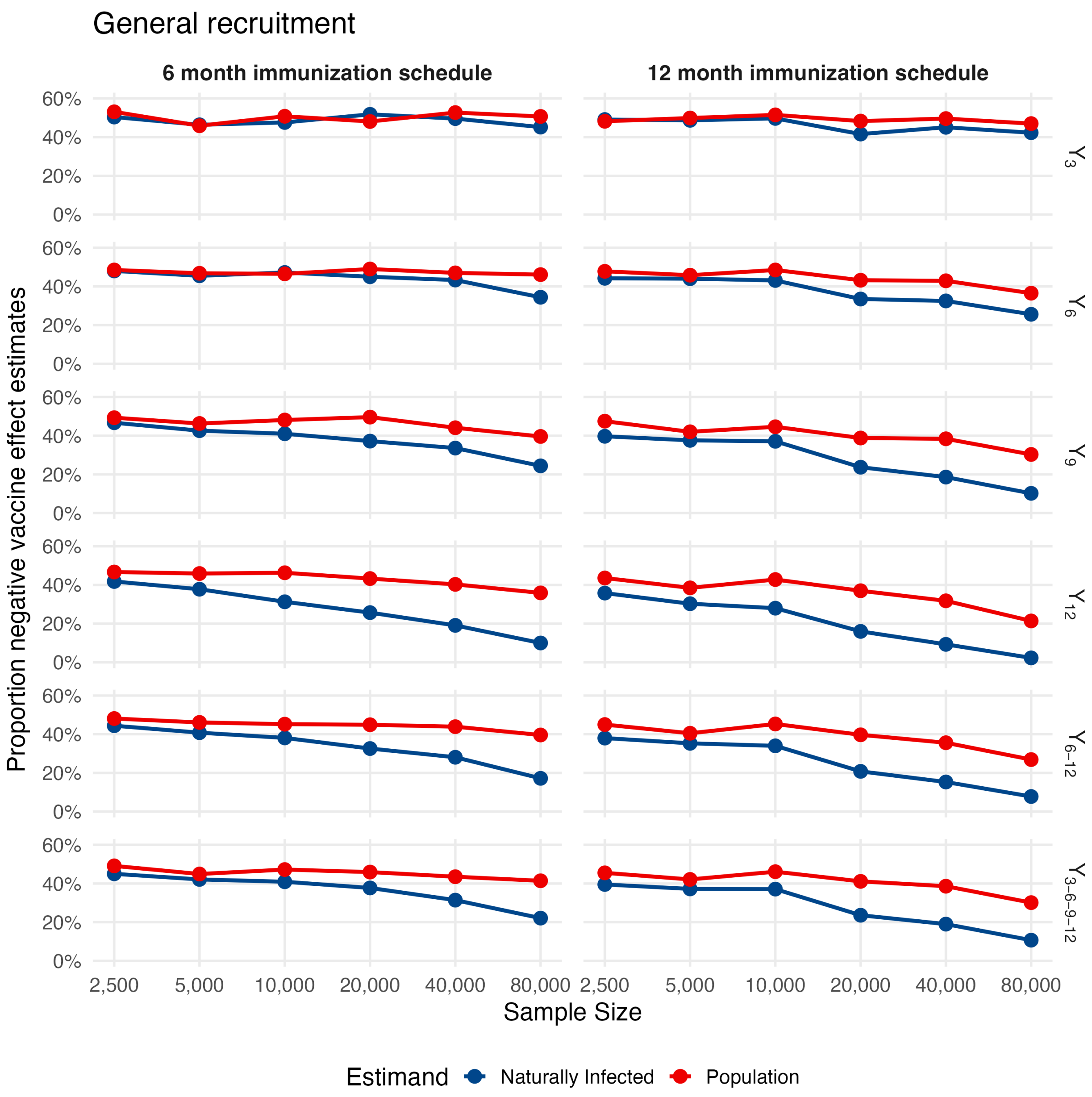

Figure 12: **Proportion of negative estimates across simulations for all endpoints under general recruitment strategy.** Shown are power estimates for the effects at each outcome time of interest, where $Y_m$ denotes height-for-age z-score measured $m$ months after infection. Columns correspond to immunization schedules with full protection at 6 or 12 months of age. Naturally infected estimates are shown in blue, while population-level estimates are shown in red.

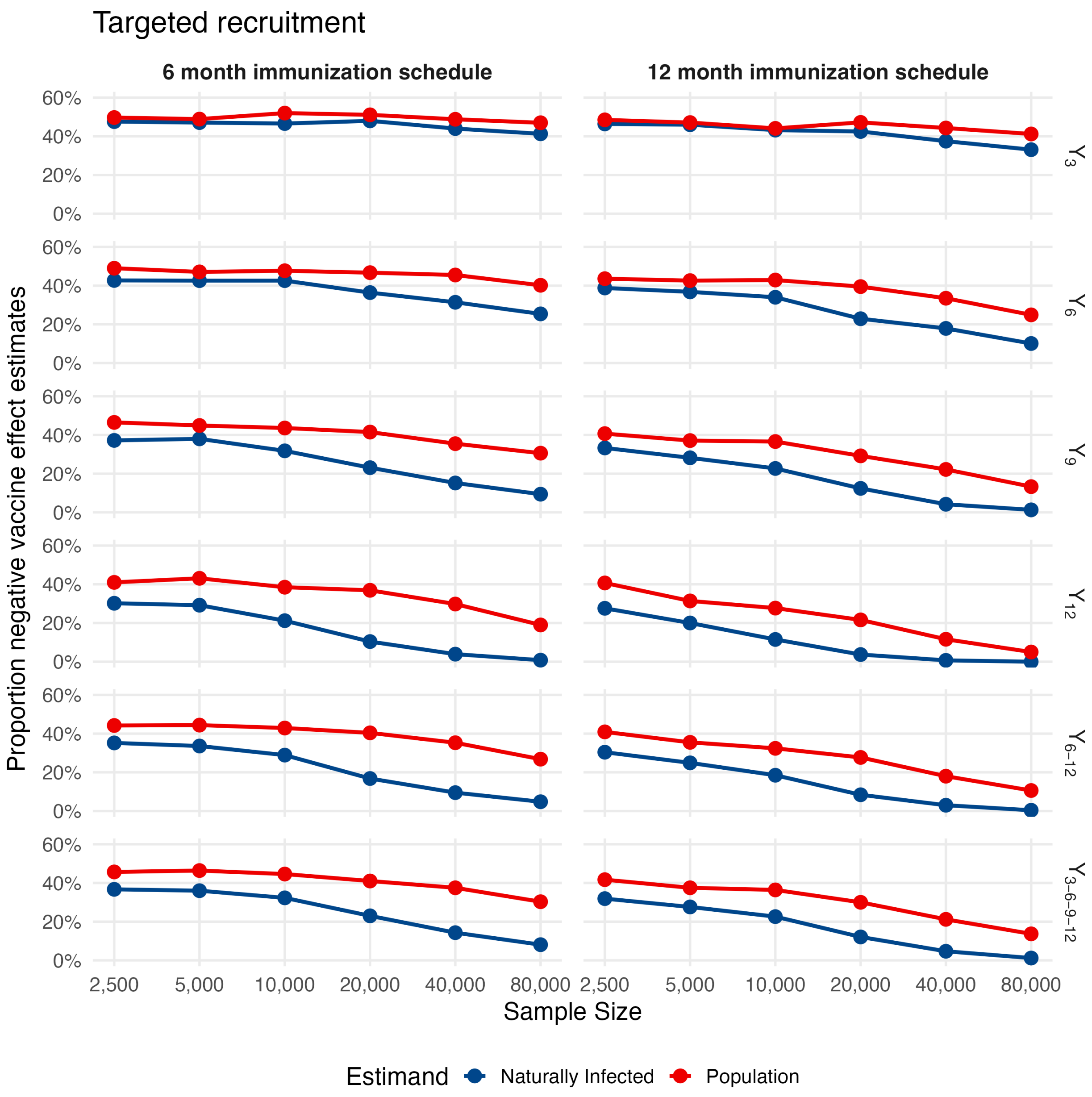

Figure 13: **Proportion of negative estimates across simulations for all endpoints under targeted recruitment strategy.** Shown are the proportion of trial simulations with negative point estimates for the effects at each outcome time of interest, where $Y_m$ denotes height-for-age z-score measured $m$ months after infection. Columns correspond to immunization schedules with full protection at 6 or 12 months of age. Naturally infected estimates are shown in blue, while population-level estimates are shown in red.

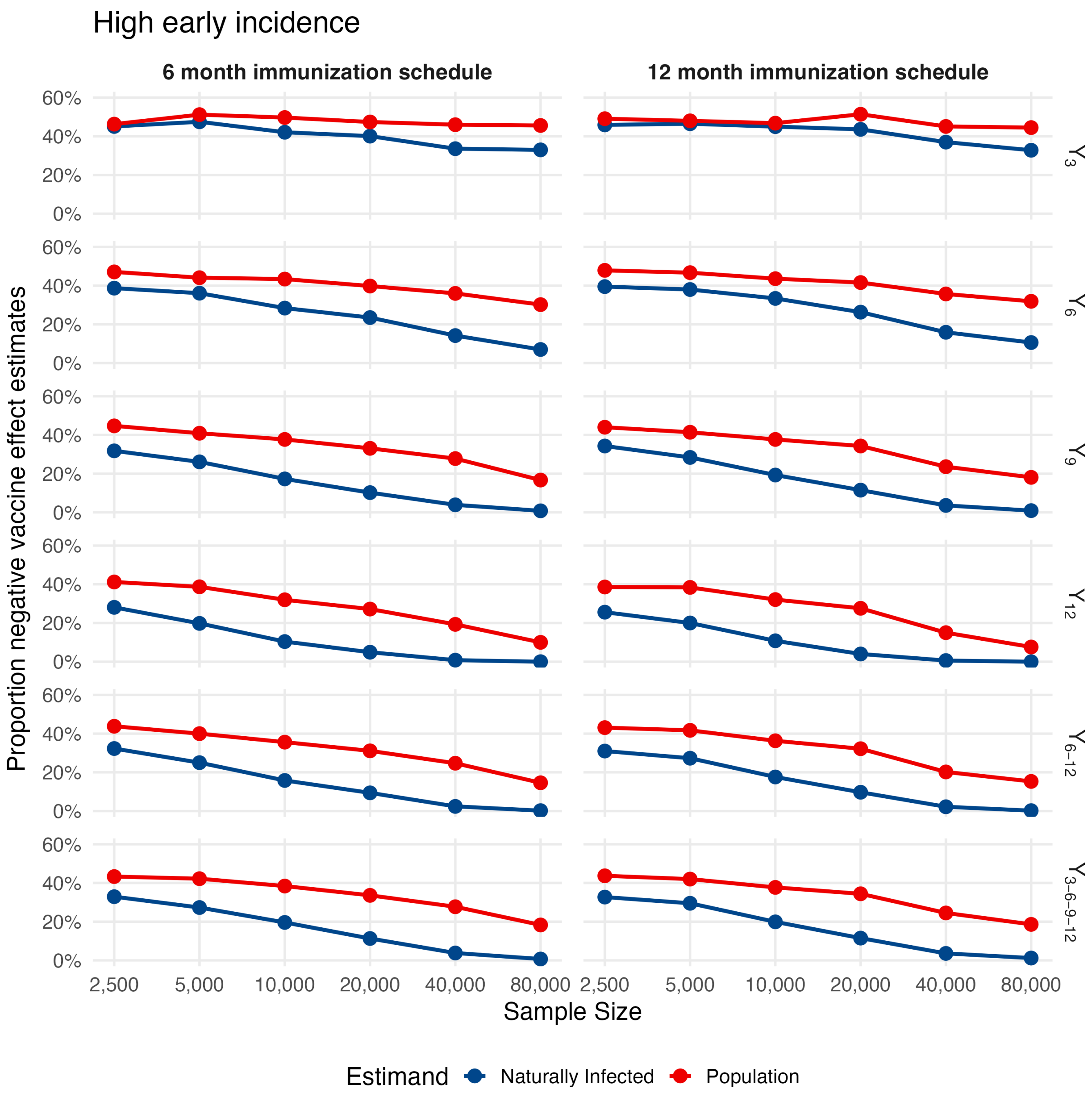

Figure 14: **Proportion of negative estimates across simulations for all endpoints under high early incidence recruitment strategy.** Shown are the proportion of trial simulations with negative point estimates for the effects at each outcome time of interest, where $Y_m$ denotes height-for-age z-score measured $m$ months after infection. Columns correspond to immunization schedules with full protection at 6 or 12 months of age. Naturally infected estimates are shown in blue, while population-level estimates are shown in red.

##### 3.4 Proportion of significant negative estimates across simulations

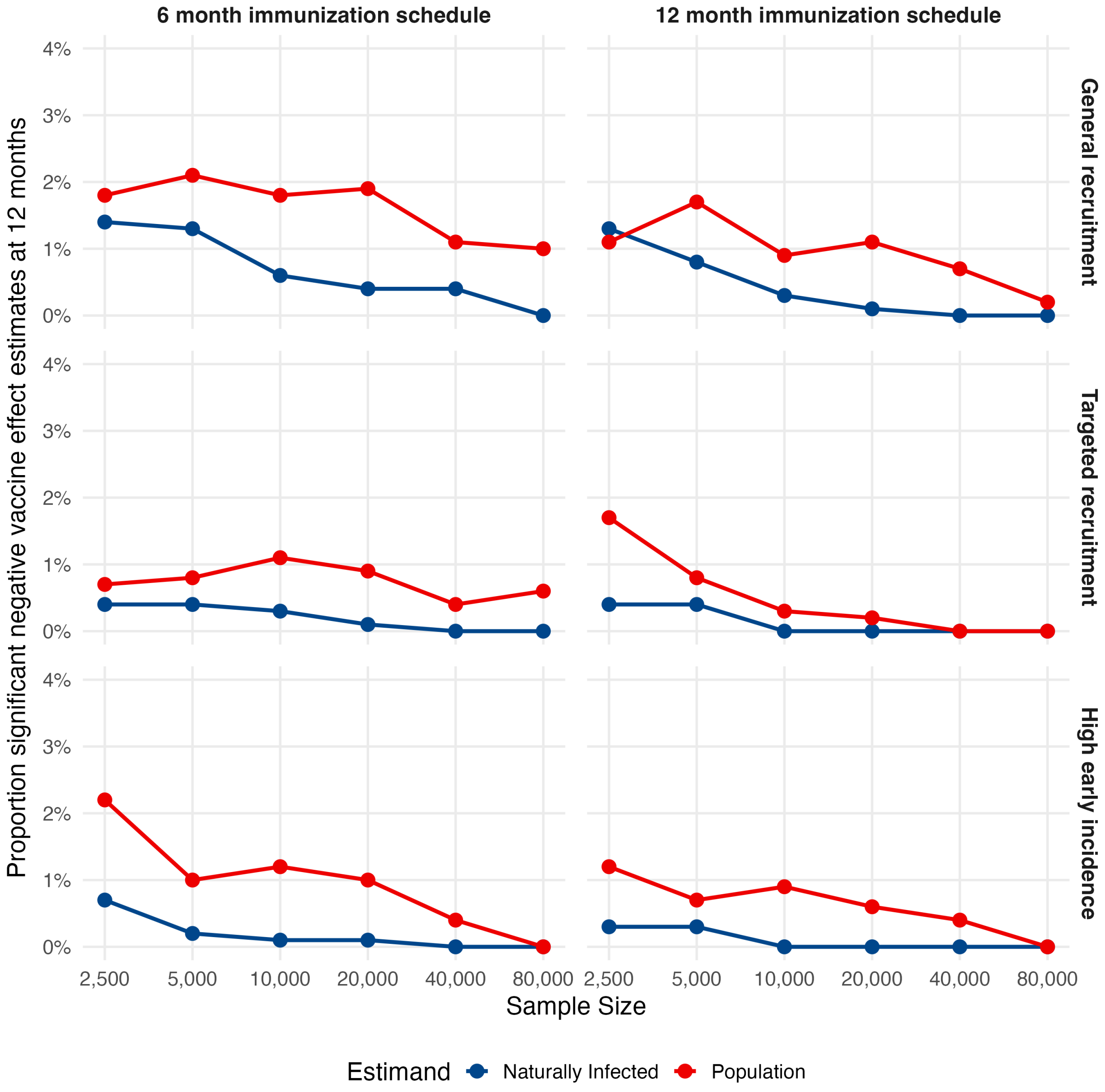

Figure 15: **Proportion of significant negative estimates across simulations for 12 month HAZ outcome across all recruitment strategies.** Columns correspond to trials using 6 month vs 12 month immunization schedules. Naturally infected estimates are shown in blue, while population-level estimates are shown in red.

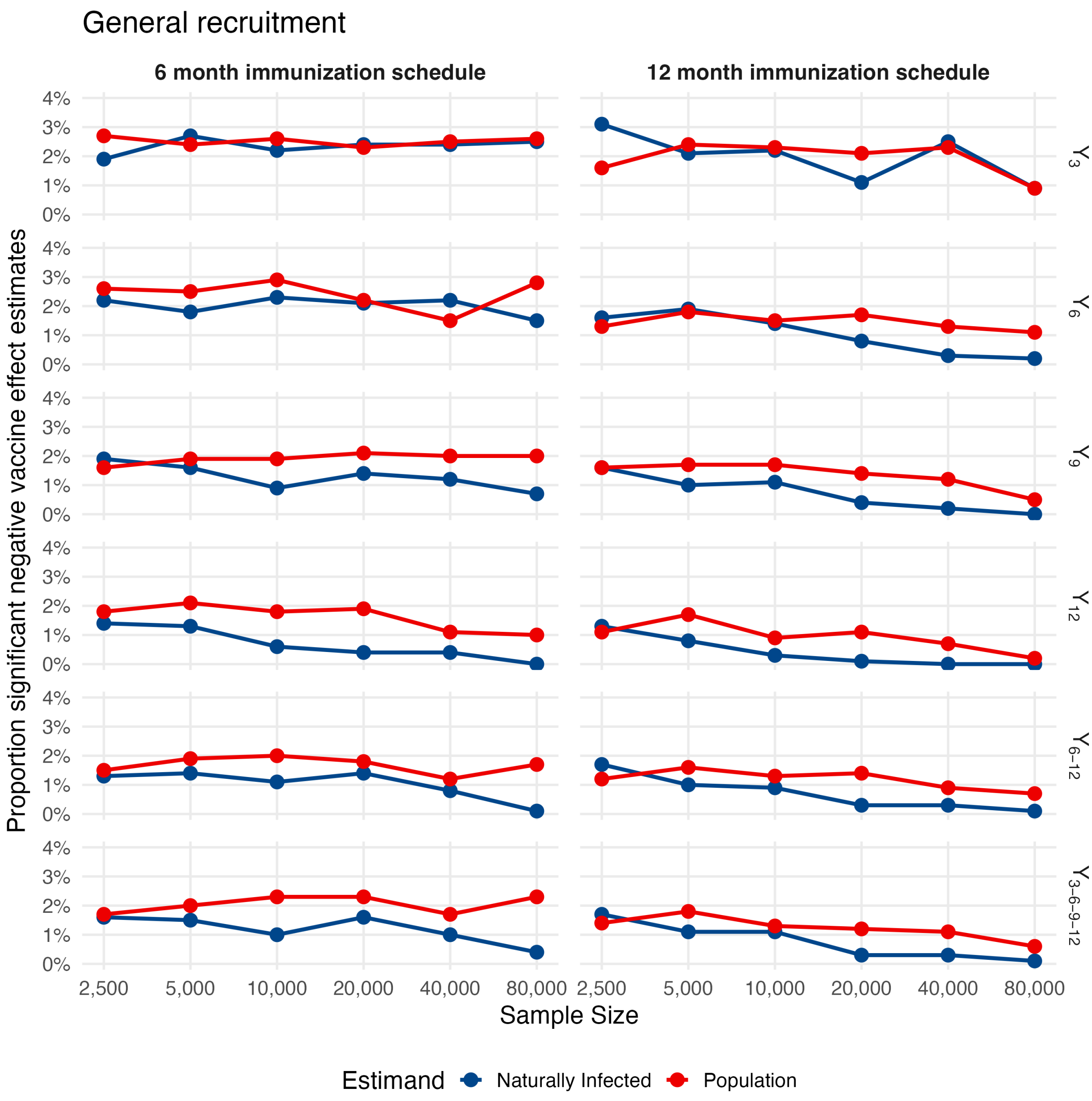

Figure 16: **Proportion of statistically significant negative estimates across simulations for all endpoints under general recruitment strategy.** Shown are the proportion of trial simulations with negative point estimates for the effects at each outcome time of interest, where $Y_m$ denotes height-for-age z-score measured $m$ months after infection. Columns correspond to immunization schedules with full protection at 6 or 12 months of age. Naturally infected estimates are shown in blue, while population-level estimates are shown in red.

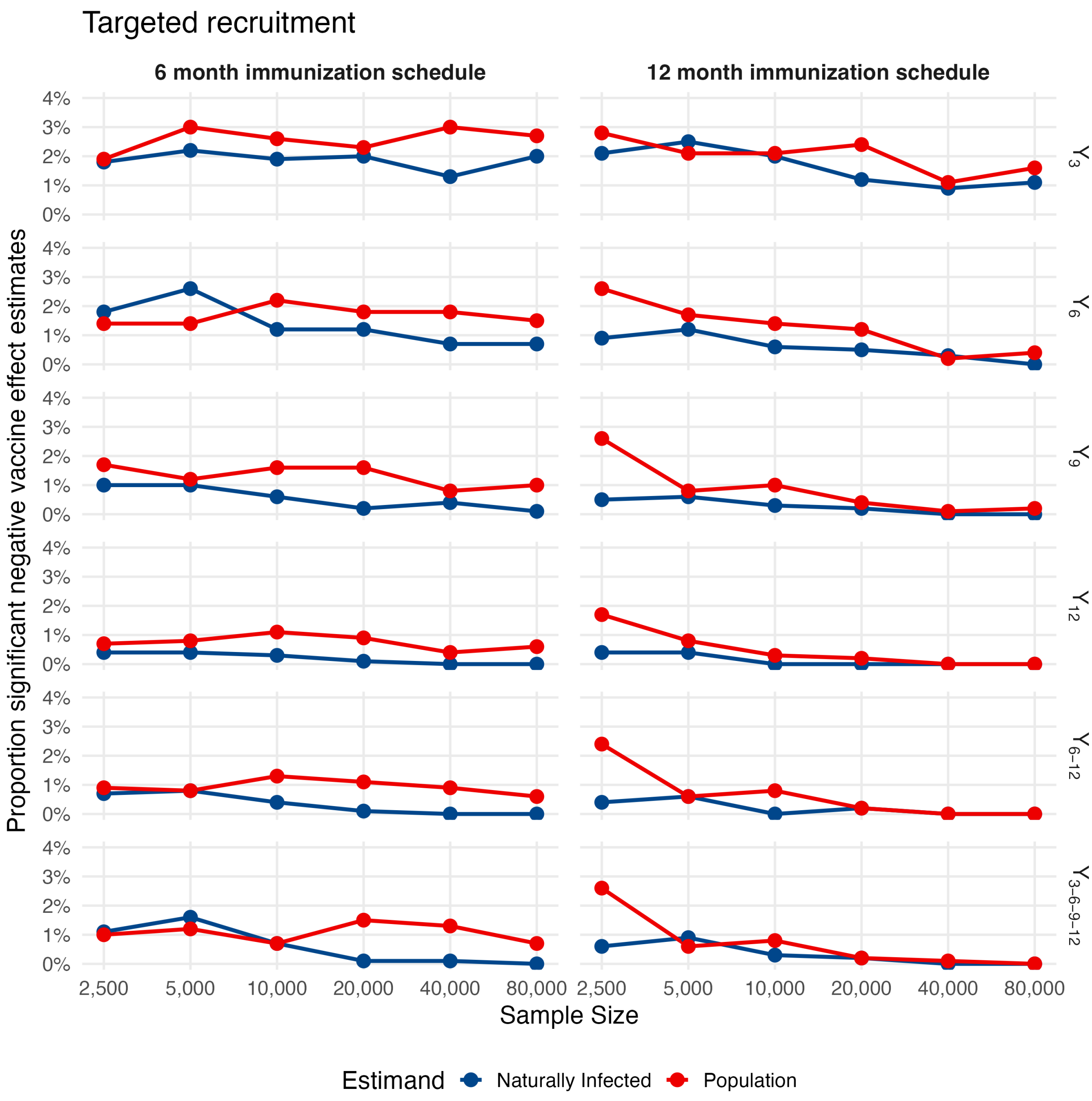

Figure 17: **Proportion of statistically significant negative estimates across simulations for all endpoints under targeted recruitment strategy.** Shown are the proportion of trial simulations with negative point estimates for the effects at each outcome time of interest, where $Y_m$ denotes height-for-age z-score measured $m$ months after infection. Columns correspond to immunization schedules with full protection at 6 or 12 months of age. Naturally infected estimates are shown in blue, while population-level estimates are shown in red.

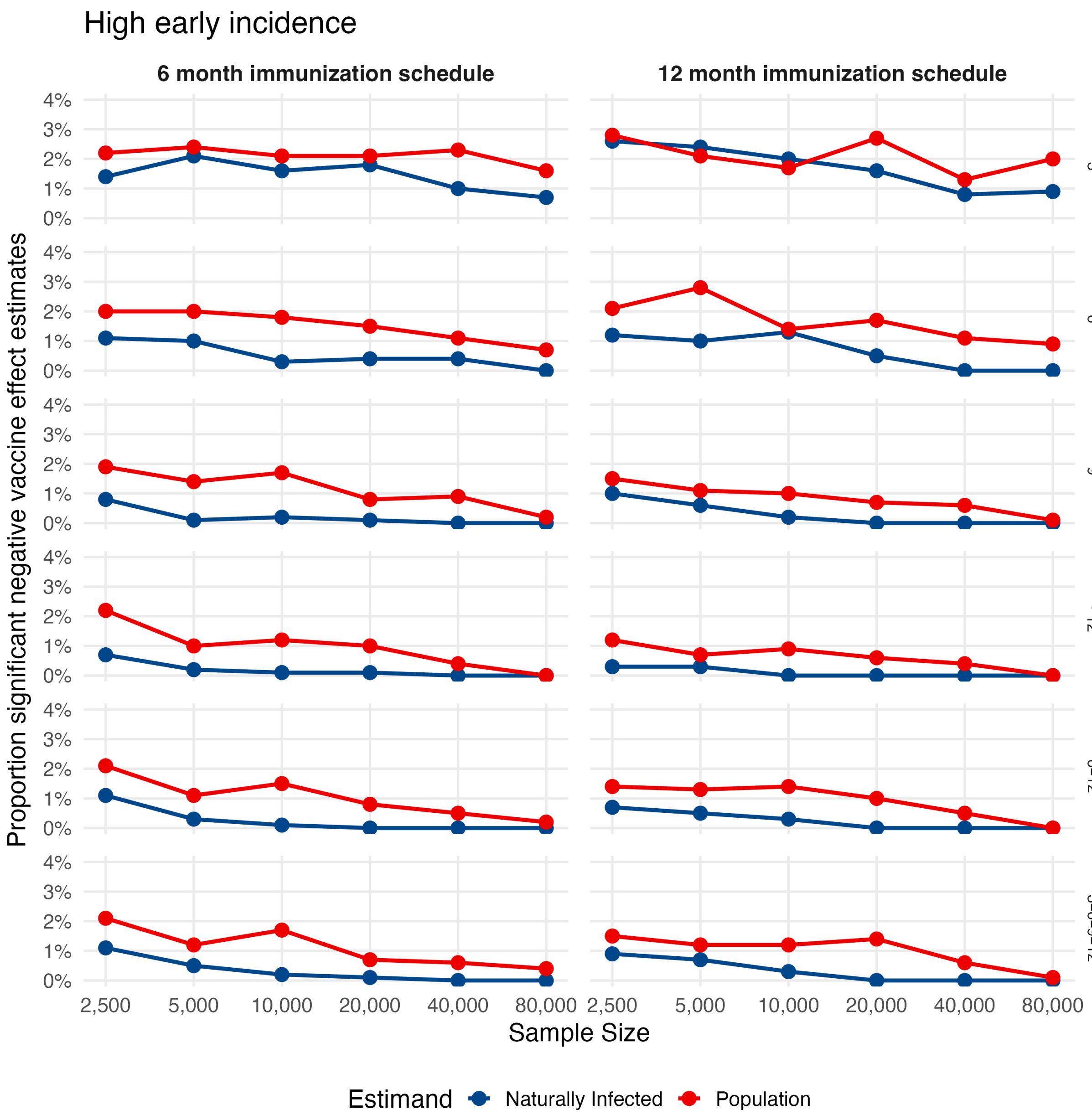

Figure 18: **Proportion of statistically significant negative estimates across simulations for all endpoints under high early incidence recruitment strategy.** Shown are the proportion of trial simulations with negative point estimates for the effects at each outcome time of interest, where $Y_m$ denotes height-for-age z-score measured $m$ months after infection. Columns correspond to immunization schedules with full protection at 6 or 12 months of age. Naturally infected estimates are shown in blue, while population-level estimates are shown in red.

[2] Hudgens MG, Hoering A, Self SG. On the analysis of viral load endpoints in HIV vaccine trials. Statistics

in Medicine. 2003;22(14):2281-98. _eprint: https://onlinelibrary.wiley.com/doi/pdf/10.1002/sim.1394. Available from: https://onlinelibrary.wiley.com/doi/abs/10.1002/sim.1394.

[3] Frangakis CE, Rubin DB. Principal Stratification in Causal Inference. Biometrics. 2002 Mar;58(1):21-9. Available from: https://doi.org/10.1111/j.0006-341X.2002.00021.x.

[4] Oxman MN, Levin MJ, Johnson G, Schmader K, Straus S, Gelb L, et al. A vaccine to prevent herpes zoster and postherpetic neuralgia in older adults. New England Journal of Medicine. 2005;352(22):2271-84.

[5] Mehrotra DV, Li X, Gilbert PB. A comparison of eight methods for the dual-endpoint evaluation of efficacy in a proof-of-concept HIV vaccine trial. Biometrics. 2006;62(3):893-900.
